# A Deep Learning Lung Cancer Segmentation Pipeline to Facilitate CT-based Radiomics

**DOI:** 10.1101/2025.06.17.25329213

**Authors:** Alfred Chung Pui So, Daryl Cheng, Shahab Aslani, Mehran Azimbagirad, Daisuke Yamada, Roberta Dunn, Eleni Josephides, Ellie McDowall, Annie-Rose Henry, Andrea Bille, Nishanth Sivarasan, Adam Pennycuick, Eleni Karapanagiotou, Joseph Jacob

## Abstract

**Background:** CT-based radio-biomarkers could provide non-invasive insights into tumour biology to risk-stratify patients. One of the limitations is laborious manual segmentation of regions-of-interest (ROI). We present a deep learning auto-segmentation pipeline for radiomic analysis.

**Patients and Methods:** 153 patients with resected stage 2A-3B non-small cell lung cancer (NSCLCs) had tumours segmented using nnU-Net with review by two clinicians. The nnU-Net was pretrained with anatomical priors in non-cancerous lungs and finetuned on NSCLCs. Three ROIs were segmented: intra-tumoural, peri-tumoural, and whole lung. 1967 features were extracted using PyRadiomics. Feature reproducibility was tested using segmentation perturbations. Features were selected using minimum-redundancy-maximum-relevance with Random Forest-recursive feature elimination nested in 500 bootstraps.

**Results:** Auto-segmentation time was ∼36 seconds/series. Mean volumetric and surface Dice-Sørensen coefficient (DSC) scores were 0.84 (±0.28), and 0.79 (±0.34) respectively. DSC were significantly correlated with tumour shape (sphericity, diameter) and location (worse with chest wall adherence), but not batch effects (e.g. contrast, reconstruction kernel). 6.5% cases had ‘missed’ segmentations; 6.5% required major changes. Pre-training on anatomical priors resulted in better segmentations compared to training on tumour-labels alone (*p*<0.001) and tumour with anatomical labels (*p*<0.001).

Most radiomic features were not reproducible following perturbations and resampling. Adding radiomic features, however, did not significantly improve the clinical model in predicting 2-year disease-free survival: AUCs 0.67 (95%CI 0.59-0.75) vs 0.63 (95%CI 0.54-0.71) respectively (*p*=0.28).

**Conclusion:** Our study demonstrates that integrating auto-segmentation into radio-biomarker discovery is feasible with high efficiency and accuracy. Whilst radiomic analysis show limited reproducibility, our auto-segmentation may allow more robust radio-biomarker analysis using deep learning features.

## 1. Introduction

Lung cancer is the most common cause of cancer-related death worldwide [1]. Average life-expectancy in patients with lung cancer in the UK is less than 1-year, mainly due to diagnosis at an advanced stage [2]. Patients are increasingly diagnosed at an earlier stage, due to several factors including introduction of targeted lung cancer screening [2]. Furthermore, the treatment landscape of early-stage non-small cell lung cancers (NSCLC) is rapidly evolving with the additional use of chemoimmunotherapy and/or targeted therapy alongside surgery [3]. Despite this, recurrence rates can remain high. With more patients diagnosed earlier and living longer, it is important to balance the risk of overtreatment and quality of life. There are no routinely used clinical biomarkers for prognostication, with TNM staging remaining the gold standard. Promising results have been reported with novel molecular tests, ctDNA, digital pathology, and artificial-intelligence (AI) based models [4–7]. Currently, the implementation of these biomarkers remains limited by repeatability, resource-allocation, and time.

Radiological biomarkers, or radio-biomarkers, are an attractive approach for clinicians because they can provide a non-invasive three-dimensional insight into cancer biology that cannot be visually evaluated by radiologists. Radiomics, a type of radio-biomarker, is a high-throughput computational method for quantitative medical imaging analysis using predefined mathematical features. NSCLC is one of the most studied oncological subtypes within radiomic research, but implementation of radiomic biomarkers has not reached clinical translation [8,9]. Previous studies have shown moderate correlation of radiomics with NSCLC prognosis, histology, actionable-genomic alterations (AGAs), and immunophenotypes [10–15]. However, radiomic features are not easily reproduced due to heterogeneity in clinical end-points, methodological transparency, image acquisition, pre-processing, segmentation quality, feature selection, and model classifier [16–18]. In addition, many radiomic studies do not incorporate robust assessments of feature stability [19].

Automatic segmentation, or auto-segmentation, has the potential to address some of these barriers including reducing manual segmentation time, avoiding observer variability, and standardising segmentation quality. To have a deployable radio-biomarker in the clinic, auto-segmentation is necessary to make these scalable. Several auto-segmentation models have already been previously published with variable performance [20–23]. However, integrating auto-segmentation within radio-biomarker studies has been less explored. Furthermore, the performance of these models will be influenced by the type of deep learning (DL) architecture used and training datasets. Using more stable regions-of-interest (ROI), such as whole lung, may be another attractive approach due to less heterogeneity in boundary definition with auto-segmentation algorithms readily available [24].

Most NSCLC-based radiomic research has been on patients receiving radiotherapy. Previous studies in resectable NSCLC have either focused only on stage 1 disease or collectively stage 1-3 [25–28]. It is important to distinguish these cohorts as resectable stage 2-3 NSCLC represents a distinct prognostic group. It is a heterogenous group with 5-year recurrence rates between 20-60%, representing a high-risk population where treatment escalation using chemotherapy with or without immunotherapy is recommended. The LACE meta-analysis demonstrated an absolute 4-5% benefit in 5-year overall survival and disease-free survival with the addition of platinum-based chemotherapy compared to surgery alone [29]. Neoadjuvant or peri-operative chemoimmunotherapy provides an additional 30-40% reduction in recurrence compared with chemotherapy, with two trials showing overall survival benefit [30,31]. Although significant breakthroughs have been made, some patients still experience early-recurrence. Current guidelines recommend 6-monthly surveillance CT scans following radical surgery for the first two years and yearly afterwards based on expert consensus [32].

Identifying early recurrence using radio-biomarkers can facilitate locoregional therapy or early introduction of systemic anti-cancer therapy (SACT) whilst the tumour-burden remains low.

In this study, we present a research pipeline integrating semi-automated DL segmentation pretrained with anatomical priors to explore robust CT-based radiomic features in resectable stage 2-3 NSCLC.

## 2. Patients and methods

This study complied with all relevant ethical regulations and received institutional approval. Informed consent was waived as this study uses a real-world database comprising routinely-collected, standard-of-care patient data and incorporates an opt-out consent process through Guy’s Cancer Cohort (Reference: 18/NW/0297) [33].

### 2.1 Patients

Patients were retrospectively identified from our tertiary cancer centre’s database from January 2015 to December 2024. All patients with resected stage 2A-3B NSCLC (TNM 9^th^ edition) were reviewed. 2-year disease-free survival (DFS) was used as the clinical endpoint. Exclusion criteria were: (1) absence of pre-operative baseline CT of the thorax, (2) follow-up time under 2-years, (3) death before recurrence, (4) inadequate information on histology and staging, and (5) pure or mixed neuroendocrine histology. 201 patients were assessed, of which 153 were eligible [Supplemental Fig. 1].

### 2.2 Image acquisition and processing

No cases were excluded based on CT acquisition protocols. 68.6% were intravenous contrast-enhanced scans and 57.5% used sharp convolution kernels. Average parameters include slice thickness 1.2mm, pixel size 0.76mm, and tube energy 113mA [Supplemental Table 1]. All images were resampled to isotropic voxel 1×1×1mm and set to lung window (level:-600HU; width: 1500HU). Images underwent grey-level normalisation 0-255 and fixed bin width discretisation to 25.

### 2.3 Image segmentation

Primary tumours were auto-segmented using nnU-Net [34]. The nnU-Net was pretrained on 23 non-cancer containing cases with anatomical priors: airway, vessel, lung [35] [Supplemental Table 2]. The model was then finetuned on 50 cancer-containing cases derived from The Cancer Imaging Archive (TCIA) [36–40] [Supplemental Table 3]. All TCIA segmentations were reviewed and corrected as necessary. The hierarchical order of label input was lung, airway, vessel, tumour. This approach to model training mirrored how a radiologist would learn, first by understanding the thoracic structures in healthy and pathological lungs, then identifying lung cancers within their anatomical context [Supplemental Fig. 2]. This type of learning, like curriculum and anatomy-aware training, has been previously described but remains under researched in lung cancers [41–42]. The nnU-Net’s default hyperparameters and 3D configuration were used.

All intra-tumoural segmentations were checked using a two-step review process and corrected on 3D-Slicer. The first review was performed by a medical oncology registrar with interest in thoracic oncology (A.C.P.S). The second review was performed by a thoracic radiologist with 10-years of experience (D.Y). Whole lungs were auto-segmented using TotalSegmentator [43]. Peri-tumoural segmentation was created by dilating the intra-tumoural segmentation by 3mm followed by removing the intra-tumoural region [Fig. 1]. Although the 3mm dilation was chosen, there is currently no standardised definition of peri-tumoural region [44].

**Figure 1:**
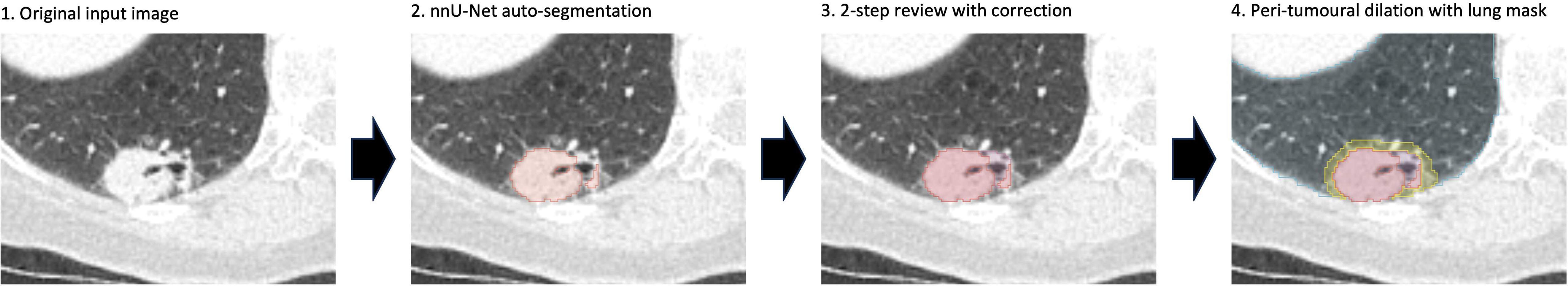
Segmentation pipeline. Both solid and non-solid components were considered as part of the segmentation. Peri-tumoural vasculature and distorted lung architecture were not included as part of the segmentation. During peri-tumoural dilation, the lung mask formed a boundary to prevent the dilation of the peri-tumoural segmentation into the thoracic wall and mediastinal structures.

### 2.4 Contour perturbations

Perturbation methods, previously shown to be comparable to test-retest studies, were used to ensure feature repeatability [45]. Five perturbations (1mm) were performed: dilation, erosion, y-axis translation, x-axis translation, and contour randomisation. Gaussian noise perturbation was not included as the cohort already have heterogenous CT acquisition parameters. Perturbation methods were not performed on whole lung segmentations as there is less variation and would result in inclusion of non-lung segments.

### 2.5 Feature extraction and processing

Radiomic features were extracted using PyRadiomics 3.1.0, an Image Biomarker Standardisation Initiative (IBSI) compliant software, and included the following filters: Laplacian of Gaussian, Wavelet, Square, SquareRoot, Logarithm, Exponential, Gradient, and LocalBinaryPattern3D [46]. A total 1967 radiomic features were extracted. Radiomic features were z-scaled.

Multivariate Imputation by Chained Equations (MICE) was used to impute missing clinical data. AGAs, ethnicity, performance status, spread-through-air-space, and PDL1 data were excluded due to high missingness. One-hot and ordinal encoding was used. Continuous variables were z-scaled.

### 2.6 Radiomic feature selection

No standardised feature selection approach exists, with each method offering different advantages and disadvantages [19]. In this study, a hybrid supervised feature selection approach of intraclass correlation coefficient (ICC) followed by minimum-redundancy-maximum-relevance recursive feature elimination-Random Forest (mRMR-RFE-RF) was used to account for feature repeatability, multicollinearity, and non-linear interactions [Fig. 2]. We used the default RF hyperparameters on sklearn and set the RFE feature minimum to 10. This entire pipeline is nested within 500 bootstrap iterations to assess for feature stability. The performance of each bootstrap is aggregated for average model performance on the hold-out test set.

**Figure 2:**
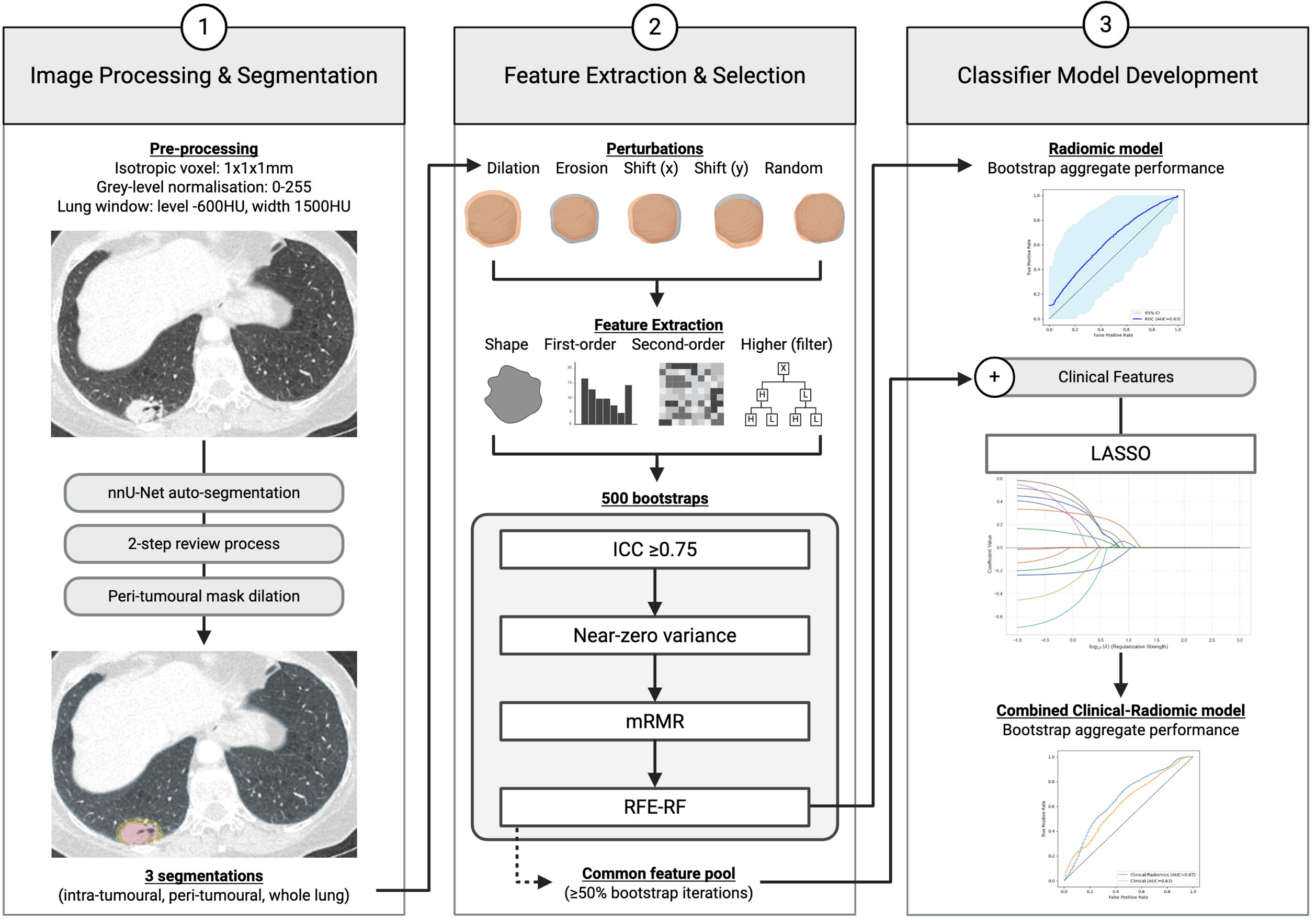
Schematic representation of the study’s pipeline. After segmentation and review, intra-tumoural and peri-tumoural segmentations underwent five perturbations: dilation (1mm), erosion (1mm), y-axis translation (1mm), x-axis translation (1mm), and contour randomisation (1mm). Features were extracted within each perturbation using PyRadiomics with the application of filters for higher-order features. Features were initially filtered based on robustness against perturbations using a conservative ICC of ≥0.75. An ICC of 0.75-0.9 and ≥0.9 indicates good and excellent repeatability respectively. Following initial filtering, features with near-zero variance were removed. Features that remained then underwent mRMR, a pairwise filter method that maximises relevance and removes redundancy through mutual information. It is used as our first step to reduce the feature pool of highly redundant features before further feature selection. A pseudo-elbow method was used which plotted the average mRMR feature importance scores against number of features added, allowing a more data-driven selection. The next step involved using RFE-RF, a feature selection wrapper method that trains a RF model and progressively removes features until the final desired number of features remain. Predictive importance of radiomic features is tested through 500 bootstrap iterations. The most common radiomic features, present across ≥50% of bootstrap iterations, are combined with clinical features and undergo further LASSO feature reduction before model building. ICC, intraclass correlation coefficient. LASSO, least absolute shrinkage and selection operator. mRMR, minimum-redundancy-maximum-relevance. RFE-RF, recursive feature elimination-Random Forest. Figure created with BioRender.

### 2.7 Clinical feature selection

The estimated maximum events per predictor (EPP) was 7 using Riley et al’s 2020 method (sample size 153, outcome proportion 0.49, R^2^_CS_ 0.15, 8 0.05) [47]. This restriction was not relevant to RF as it is less reliant on the traditional EPP-based restrictions. Clinical features were selected through a combination of clinical features of interest, assessment of multicollinearity, and backward feature elimination. The selected features were trained using logistic regression across 500 bootstraps.

### 2.8 Combined clinical-radiomic model

Combining clinical with radiomic features can be done at an early or late stage of a feature selection pipeline. Although early fusion is advantageous as it considers interactions between clinical and radiomic features early on, RF is often biased towards selecting continuous over categorical features based on how it calculates feature importance due to splitting opportunities [48]. To address this, clinical and radiomic features were combined after their individual selection process. To limit the number of features utilised (EPP=7), we used least absolute shrinkage and selection operator (LASSO) to reduce the feature pool.

### 2.8 Statistical analysis

The performance of the auto-segmentation model was evaluated using volumetric DSC (vDSC), Intersection over Union (IoU), sensitivity, specificity, precision, and accuracy. Hausdorff distance was not calculated as it can be confounded by presence of synchronous lesions. Surface DSC (sDSC) was reported as it more closely predicts time-saved in manual segmentation compared to overlap metrics [49]. A 1mm sDSC tolerance threshold was used to reflect a strict threshold within the maximum boundary variation introduced by perturbations. The final segmentations used within the prediction model was considered the ground-truth as it had been reviewed by the thoracic radiologist who has been blinded to this study.

Spearman rank and Mann-Whitney U were used to correlate independent variables with DSC scores. Kruskal-Wallis followed by Dunn’s post-hoc test was used for multiple comparisons between tumour location and DSC scores. Beta-regression was used for multivariate analysis in DSC score evaluation. Wilcoxon-rank was used to compare different auto-segmentation performance. Chi-squared and Mann-Whitney U were used to compare patient characteristics. The performance of the predictive model was evaluated by Receiver-Operator Characteristic (ROC) curves: Area Under the Curve (AUC), F1 score, specificity, sensitivity, recall, precision, accuracy, and brier calibration score. Probability threshold for classification was kept at default 0.5. ROC curves were compared using DeLong’s test. As this is an exploratory study, the alpha significance threshold was kept at 0.05.

All analysis was performed on Python 3.10.12: matplotlib 3.10.0, MLstatkit 0.1.7, mrmr_selection 0.2.8, nibabel 5.3.2, numpy 2.1.3, pingouin 0.5.5, pyroc 0.1.1, scikit-learn 1.6.1, scikit-optimize 0.10.2, scipy 1.15.1, SimpleITK 2.4.1, statsmodels 0.14.4, and surface-distance 0.1.

## 3. Results

### 3.1 Patient characteristics

153 patients were included in this study with a median age of 69 years old (range 46-88), 43.8% male sex, 75.8% non-squamous cell histology, and 56.2% stage 3. 88.2% of patients had a lobectomy. 7.2% and 50.3% of patients received neoadjuvant and adjuvant SACT respectively [Table 1]. The 2-year DFS rate following surgical treatment was 49.0% (*n*=75/153). Of those that recurred within 2 years, the median time-to-recurrence was 252 days (95%CI 221-325 days) [Supplemental Fig. 3].

**Table 1:**
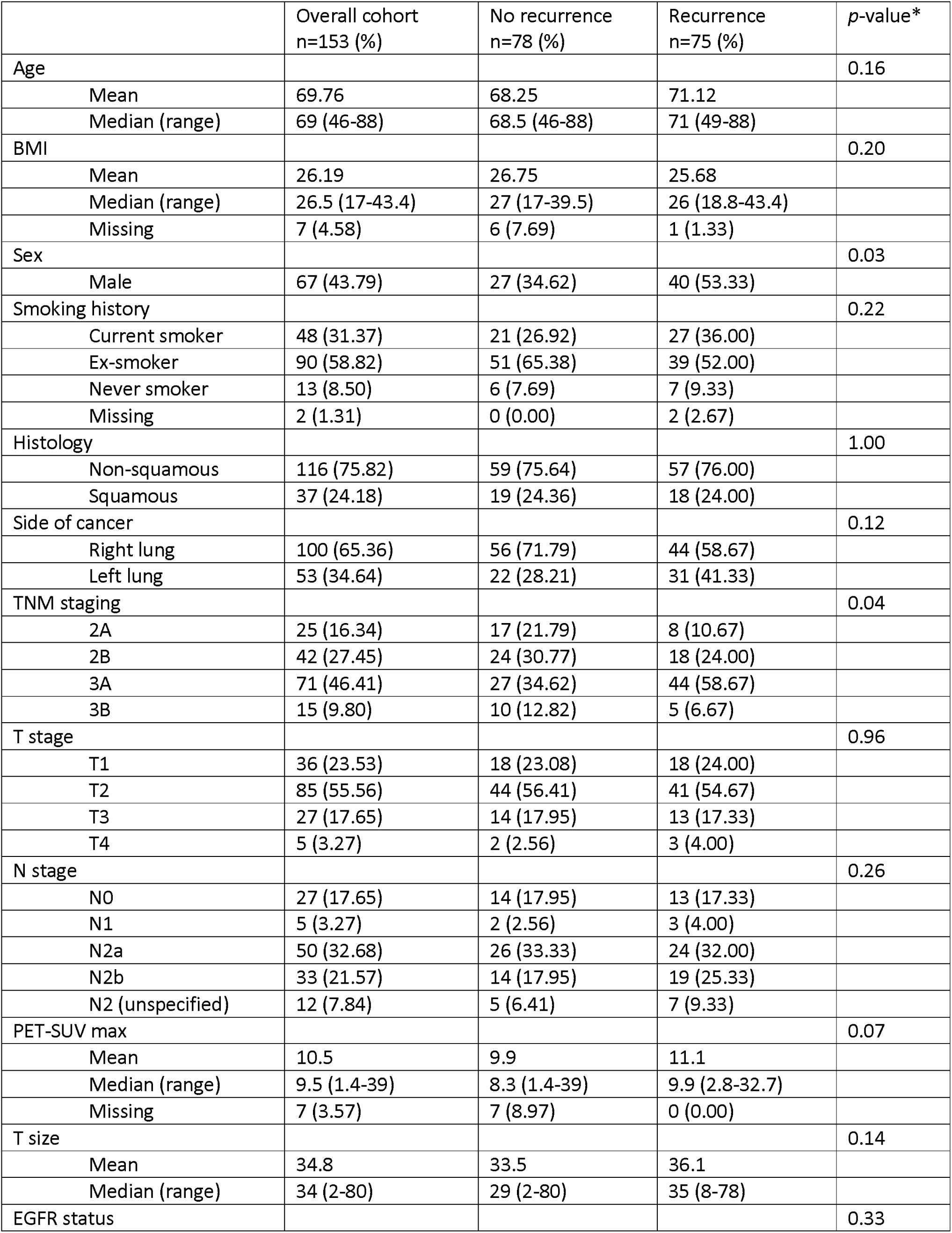
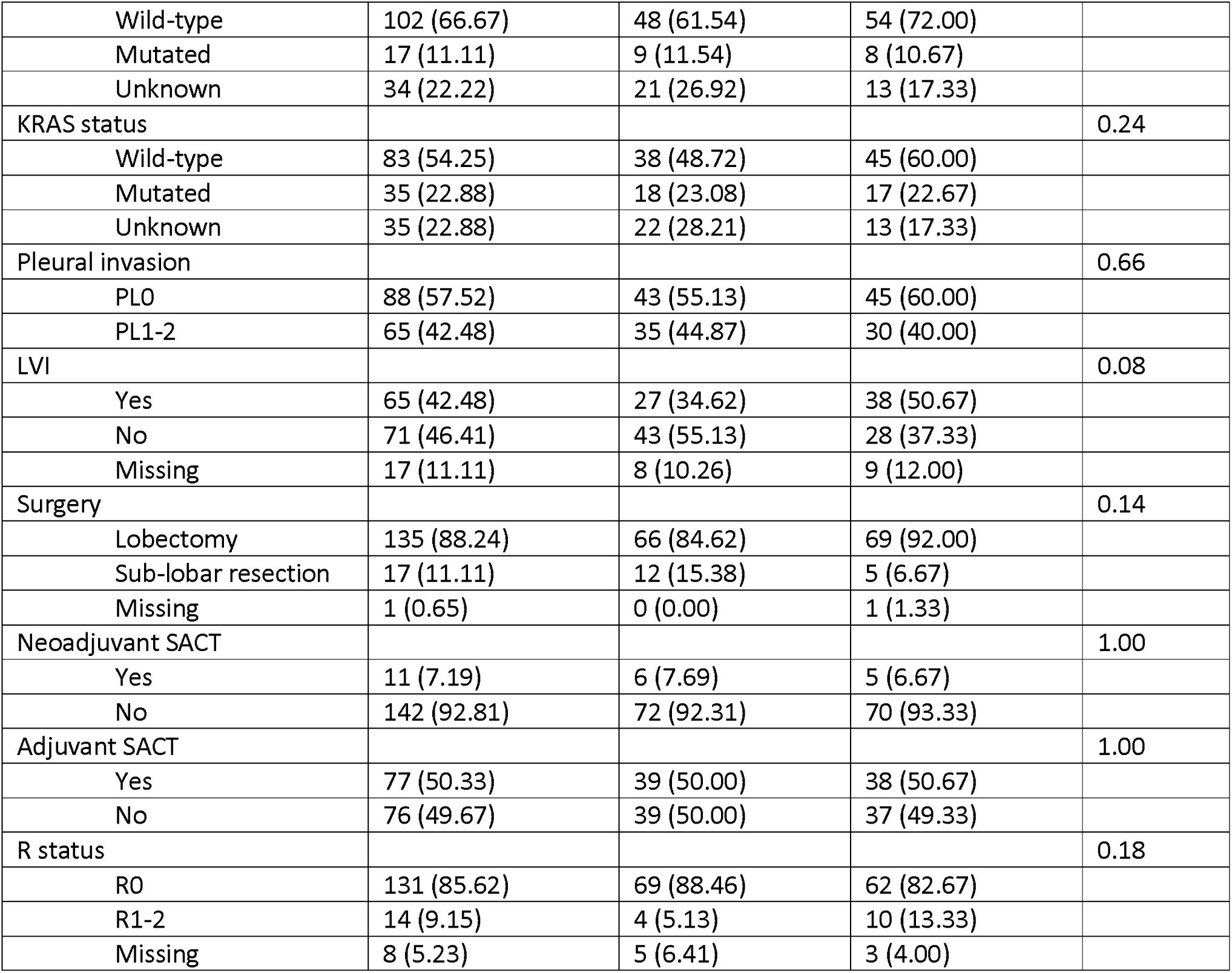
Patient baseline demographics, cancer-specific characteristics, and treatment regimens received by patients. Percentages are displayed as within column percentages. The p-value was calculated using Chi-squared test for categorical variables and Mann-Whitney U test for continuous variables.

### 3.2 Auto-segmentation performance

The average auto-segmentation time per patient case was 36 seconds (range 12-91), equating to ∼7.7 slices per second. This includes segmenting the tumour, whole lungs, airways, and vasculature. Our model had good performance across all metrics with mean vDSC, sDSC, IoU, sensitivity, specificity, precision, and recall of 0.84 (±0.28), 0.79 (±0.34), 0.80 (±0.31), 0.89 (±0.28), 1.00 (±<0.01), 0.86 (±0.26), and 1.00 (±<0.01) respectively [Fig. 3]. In total, there were 6.5% (n=10/153) ‘missed’ segmentations and 6.5% (n=10/153) segmentations that required major changes. Majority of these tumours were adherent to extra-pulmonary structures or had parenchymal distortion [Supplemental Fig. 4, Supplemental Table 4].

**Figure 3:**
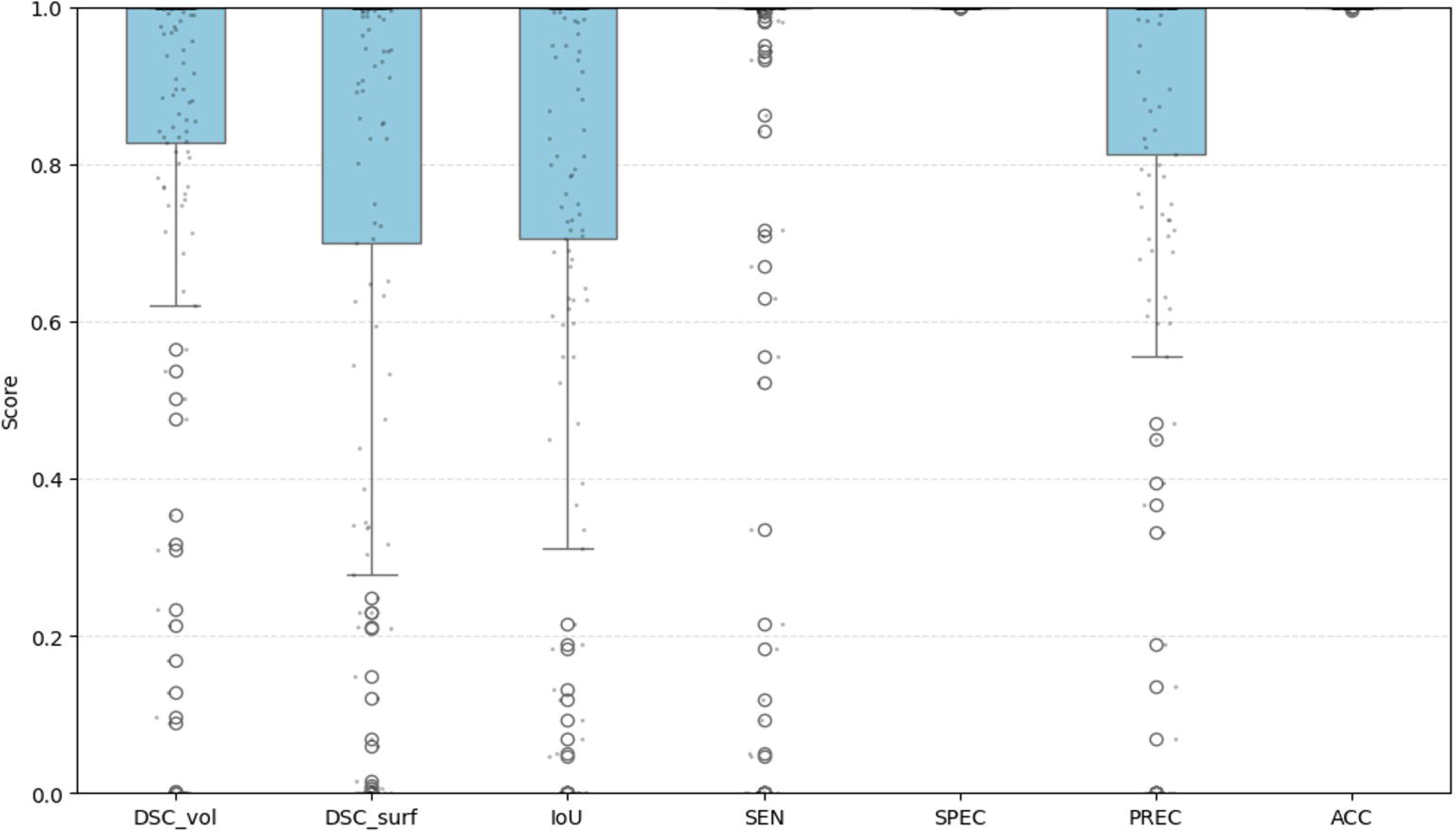
Box-whiskers plot reporting standard metrics for the pre-trained auto-segmentation model (model_A). ACC, accuracy. DSC, Dice-Sørensen coefficient. DSC_surf, surface DSC. DSC_vol, volumetric DSC. IoU, Intersetion over Union. PREC, precision. SEN, sensitivity. SPEC, specificity.

### 3.3 Tumour and Batch factors on auto-segmentation performance

Six tumour factors (volume, sphericity, maximum diameter, surface:volume ratio, semi-solid component, location) and four batch factors (contrast, original slice thickness, original pixel size, kernel type) were assessed against segmentation performance. Median tumour volume and maximum 3D diameter were 10727mm^3^ (IQR 3152-21140) and 49.4mm (IQR 30.0-75.0) respectively. 20.9% and 13.7% of tumours were adhering to the chest wall and mediastinum respectively [Supplemental Table 5]. 82.4% were pure solid tumours.

On univariate analysis, both DSC scores were positively correlated with sphericity, pure solid tumours, smooth kernel, and presence of intravenous contrast [Supplemental Table 6]. Larger tumour volume and diameter had a weak inverse relationship with performance. On Dunn’s multiple comparisons, vDSC and sDSC scores were worse with tumours adhering to the hilum (*p*=0.003; *p*=0.001), apices (*p*=0.028; *p*=0.006), and chest wall (*p*=0.048; *p*=0.16) [Supplemental Fig. 5]. On multivariate beta regression, sphericity, maximum 3D diameter, and peripheral invasion (i.e. tumour adherence to chest wall, apices, diaphragm) remained independently correlated with performance [Table 2]. Involvement of the hilum or mediastinum were trending towards worse segmentation but did not reach statistical significance. Although segmentation of semi-solid nodules was comparable to pure solid nodules, visual inspection on the poorly segmented semi-solid nodules (DSC <0.6) showed that the non-solid component was commonly missed [Supplemental Fig. 4]. None of the batch factors were independently correlated with segmentation performance. All variables had a variance inflation factor (VIF) <2.

**Table 2:**
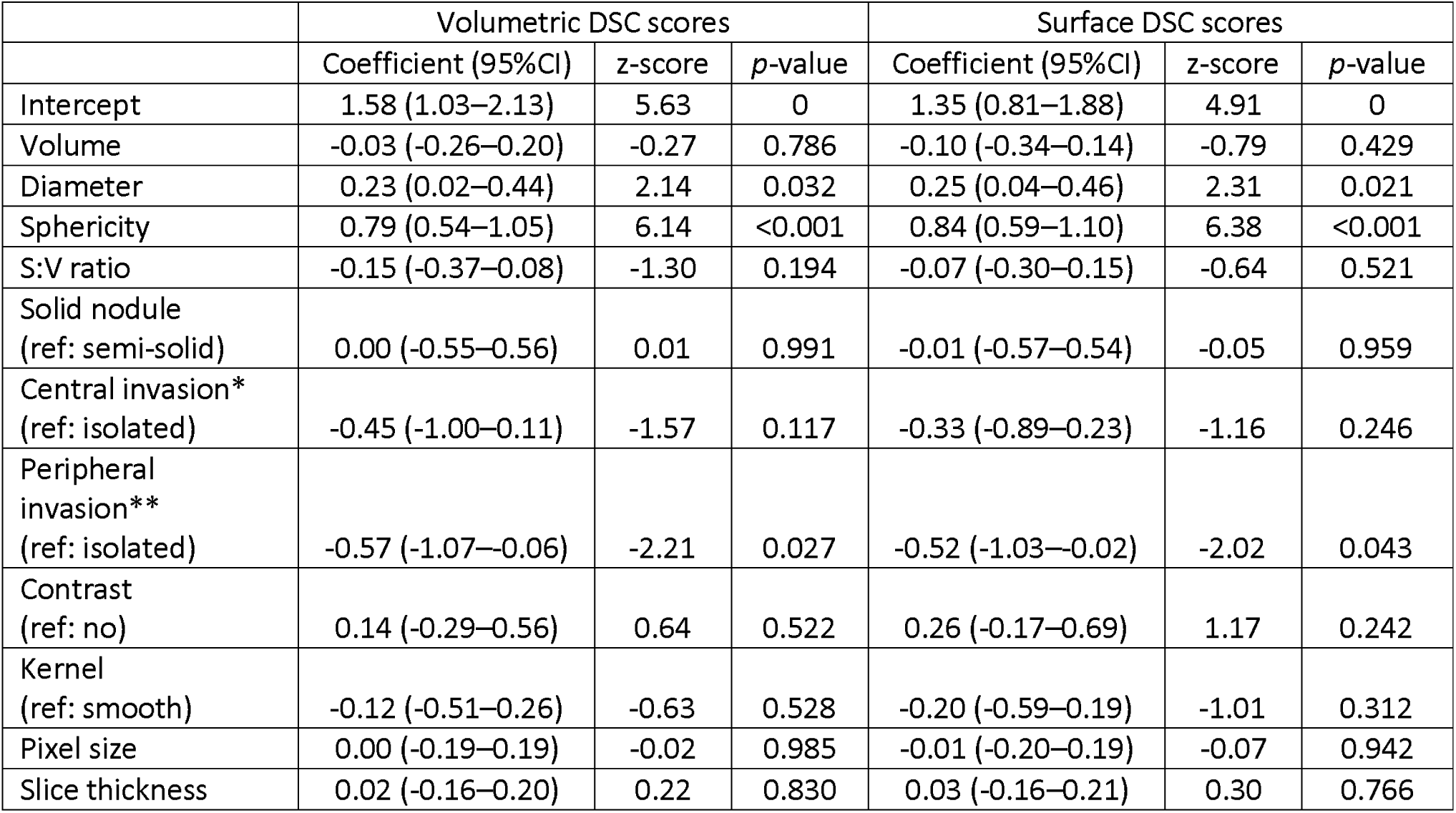
Multivariate beta regression analysis of factors impacting volumetric and surface DSC scores. Multiple linear regression (e.g. ordinary least squares) was not appropriate due to bounded outcome (DSC score 0-1), asymmetric distribution, and heteroskedasticity. Sphericity measures the roundness of the tumour region relative to a sphere, and surface:volume ratio measures the general compactness of the shape relative to the volume (a lower score indicates more sphere-like shape). Pixel size and slice thickness represent original sizes before pre-processing (all images were resampled to isotropic voxel 1×1×1mm). Isolated tumours refer to tumours not in contact with other major boundary structures (e.g. chest wall, hilum, mediastinum). Volumetric DSC: Pseudo R^2^ 0.320. Surface DSC: Pseudo R^2^ 0.348. S:V ratio, surface:volume ratio. *Central invasion refers to lesion adhering to the mediastinum (n=21/153) or hilum (n=10/153). ** Peripheral invasion refers to lesion adhering to the chest wall (n=32/153), apices (n=7/153), or diaphragm (n=5/153).

### 3.4 Dataset influence on auto-segmentation performance

To test the influence of pre-training and anatomical labels on model performance, we compared the pre-trained model (model_A) against nnU-Net models trained only on tumour-labels (model_B) and tumour-airway-vessel-lung labels (model_C) with the same TCIA dataset. Model_A performed significantly better than model_B and _C across vDSC, sDSC, IoU score, and sensitivity (*p*<0.001) [Table 3; Supplemental Fig. 6]. The addition of anatomical labels with tumour labels during training (model_C) improved all metrics scores compared to training with tumour labels alone (model_B), but only accuracy and precision reached statistical significance. Compared to both models, model_A had less missed cases and poorly segmented cases (vDSC <0.6). Interestingly, not all poorly segmented cases overlapped between models [Supplemental Fig. 7].

**Table 3:**
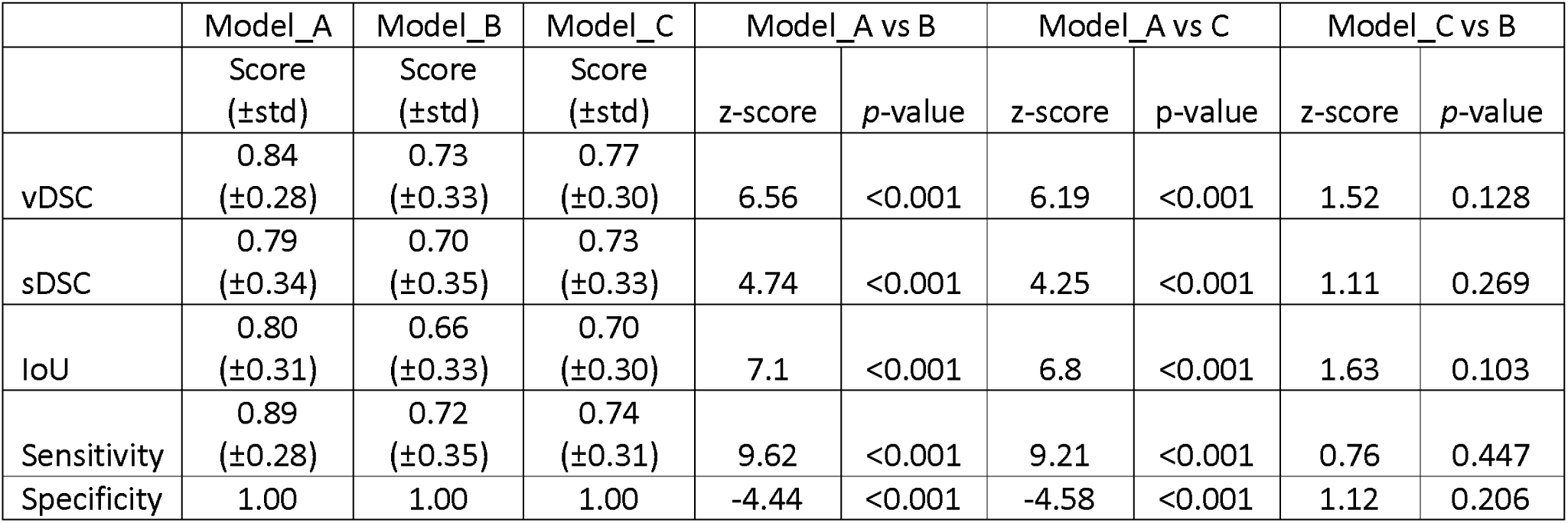
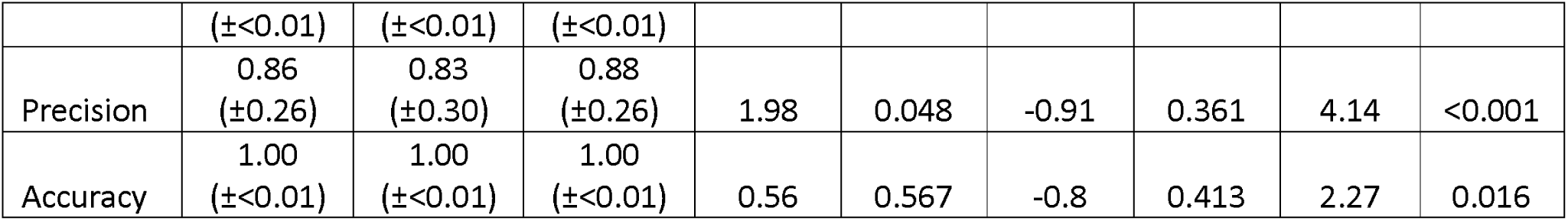
Wilcoxon-rank pairwise comparison of auto-segmentation metrics across models. Model_A: pre-trained using anatomical priors (lung, airway, vessel) on cases without lung tumours, then finetuned on cases with lung cancers. Model_B: no pre-training, only trained on tumour labels. Model_C: no pre-training, trained on tumour labels as well as anatomical labels (lung, airway, vessel). IoU, intersection over union. sDSC, surface DSC. std, standard deviation. vDSC, volumetric DSC.

On reverse validation, we employed the same transfer learning approach with anatomical priors using the GCC dataset (n=153) and validating on the unseen TCIA dataset (n=50). The pre-trained model (model_D) performed marginally better than the model trained on tumour labels alone (model_E) across all metrics but did not reach statistical significance [Supplemental Table 7; Supplemental Fig. 8].

### 3.5 Correlation of clinical features with 2-year DFS

For the clinical model, 7 clinical features were selected: staging, R status, sex, PET-SUV-max of primary tumour, reception of SACT, pleural invasion, and type of surgical treatment [Supplemental Table 8]. On multivariate analysis, only staging and sex were significantly correlated with 2-year DFS. Interaction analysis demonstrated that reception of SACT in stage 3 disease reduced probability of recurrence from 0.72 to 0.52 (marginal effect *dy/dx* = –0.32; *p*=0.076). LASSO feature reduction similarly identified staging, sex, and PET-SUV-max as prognostic features [Supplemental Fig. 9]. The multivariate model, either using features from the classic approach or LASSO, had similar predictive performance with AUCs 0.63 (95%CI 0.54-0.71) and 0.61 (95%CI 0.52-0.70) respectively [Supplemental Table 9; Supplemental Fig. 10]. Using a RF classifier did not improve performance, suggesting absence of significant non-linear interactions.

### 3.6 Intra-tumour, peri-tumoural, and whole lung radiomic models

On average, 55-60% and 20-25% of radiomic features achieved an ICC ≥0.75 and ≥0.90 across multiple perturbations. Although 150-200 features contribute >50% of total feature relevance score during mRMR, selecting the top 100 features achieved the most consistent performance. Median number of features selected across bootstraps following mRMR-RFE-RF for intra-tumoural, peri-tumoural, and whole lung radiomics were 61 (IQR 32-83), 56 (IQR 30-80), and 50 (IQR 34-78) respectively. Intra-tumoural, peri-tumoural, and whole lung radiomics had wide confidence intervals, suggesting radiomic feature instability, with AUCs 0.63 (95%CI 0.38-0.84), 0.56 (0.31-0.77), and 0.58 (95%CI 0.34-0.78) respectively [Supplemental Fig. 11]. Limiting feature count between 10-30 did not improve AUC scores. Nine intra-tumoural, 7 peri-tumoural, and 2 whole lung features were present in ≥50% of bootstrap iterations [Supplemental Table 10].

### 3.7 Clinical-radiomics model

Adding radiomic features from a single ROI source did not improve predictive performance. Combining the common intra-tumoural, peri-tumoural, and whole lung features with the clinical model followed by LASSO feature reduction improved overall performance with AUC of 0.67 (95%CI 0.59-0.75) and Brier score of 0.23 (95%CI 0.21-0.25) [Table 4, Supplemental Fig. 12]. However, this was not statistically significant compared to the clinical model (ΔAUC 0.04; z-statistic 1.08, *p*=0.281). The use of an RF classifier did not improve model performance.

**Table 4:**
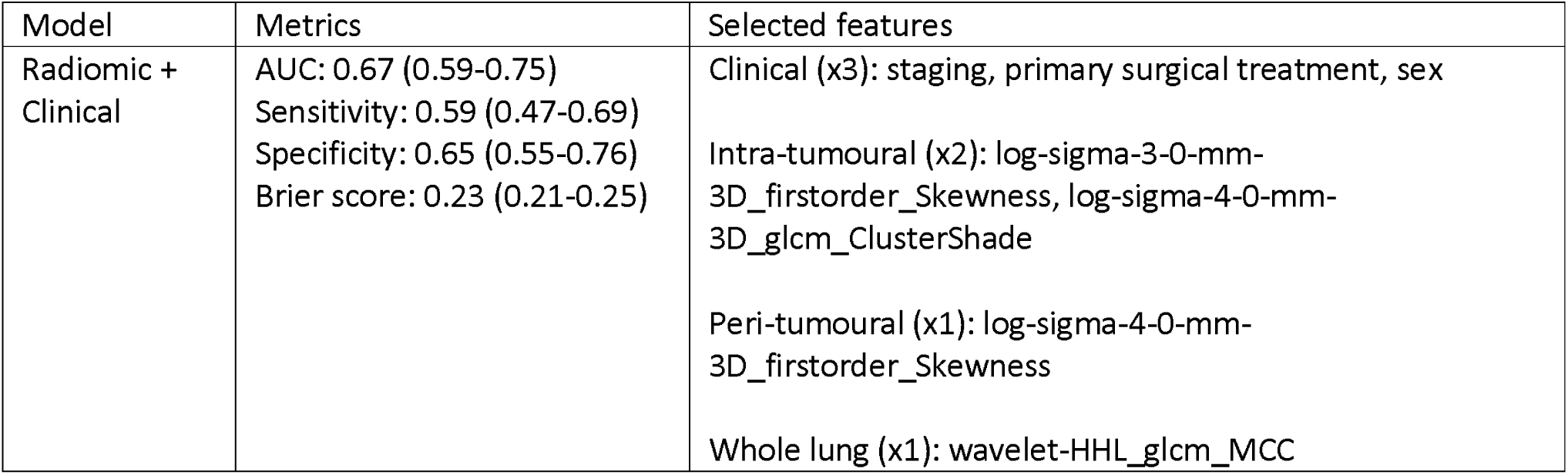
Clinical-radiomic model performance and selected feature combinations. Coefficients for the logistic regression model were: staging (3>2; +0.46), primary surgical treatment (sub-lobar resection>lobectomy;-0.49), sex (male>female; +0.51), tumour_log-sigma-3-0-mm-3D_firstorder_Skewness (-0.62), tumour_log-sigma-4-0-mm-3D_glcm_ClusterShade (-0.81), peri_log-sigma-4-0-mm-3D_firstorder_Skewness (+0.53), lung_wavelet-HHL_glcm_MCC (-0.33).

### 3.8 Interpreting radiomic features

Intra-tumoural ‘skewness’ and ‘cluster shade’ were inversely correlated with recurrence, whereas peri-tumoural ‘skewness’ was positively correlated with recurrence. ‘Skewness’ is a first order feature that represents asymmetry of the distribution of intensity along the global mean, whilst ‘cluster shade’ is a textural feature that represents asymmetry or non-uniformity of co-occurring pixels. Pearson correlation demonstrated that none of the commonly selected radiomic features have a strong correlation with tumour size, PET-SUV-max, presence of lvi, pleural invasion, and histology. Exploratory analysis suggests a moderate positive correlation of predominant micropapillary growth pattern with intra-tumoural ‘cluster shade’ (*r*=0.46-0.47; *p*<0.001) [Supplemental Fig. 12].

## 4.0 Discussion

This study demonstrated that a semi-automated DL segmentation pipeline for radio-biomarker discovery is feasible with high efficiency and good segmentation performance. The addition of pre-training using anatomical priors improved auto-segmentation performance compared to training with tumour labels alone and training with anatomical labels (without pretraining). The accuracy of the nnU-Net model was not influenced by batch effects (e.g contrast, reconstruction kernel) but rather by tumour shape and location. In general, more irregularly shaped tumours and tumours adhering to the chest wall had lower DSC scores. This study also showed that within this cohort of resected stage 2A-3B NSCLCs, CT-based traditional handcrafted radiomic features were highly unstable with poor repeatability.

In recent years, there has been growing interest in using auto-segmentation in radio-biomarker studies. This is in part due to reduction in manual segmentation time as well as enhancing consistency and reproducibility.

Furthermore, auto-segmentation provides the opportunity for radio-biomarkers to be deployable in clinical practice due to scalability. Several auto-segmentation models have been published with moderate to good performance [20–23]. For example, Primakov et al (2022) used a modified 2D CNN trained on nearly 1000 CT scans and achieved a vDSC and sDSC of 0.82±0.17 and 0.63±0.28 respectively [20]. In that study, higher tumour complexity, as measured by the necessity of PET-guided manual segmentation or tumour adherence to mediastinum or chest wall, were associated with poorer segmentations [20]. The nnU-Net is an attractive approach to segmentation as it is self-configuring and easy to adapt. Currently, evidence in integrating auto-segmentation within radio-biomarker development in lung cancer remains limited. Furthermore, surface agreement (e.g. sDSC), rather than overlap (e.g. vDSC, IoU), is more representative of time-saved from contouring but is usually not reported [49]. Our auto-segmentation model has comparable, if not better, performance compared to most studies with vDSC, sDSC, and IoU of 0.84 (±0.28), 0.79 (±0.33), and 0.80 (±0.31). Furthermore, this was achieved within stage 2-3 NSCLCs of various radiological phenotypes and CT acquisition parameters. The reason segmentation performance was not affected by batch effects was likely due to the heterogenous training dataset and nnU-Net’s self-calibration during training. The segmentations that were missed or required major revisions correlated with more irregular tumour shapes and adherence to the chest wall. Unfortunately, it is difficult to train complex tumour shapes as there is no clear agreement amongst expert clinicians regarding ground-truth boundaries [50]. On the other hand, there may be several approaches to improve auto-segmentation of tumours adhering to major boundary structures. These include increasing anatomical priors (e.g. chest wall, heart), excluding pixels in non-lung regions, limiting the input volume by cropping around the tumour region, and adding boundary streams to highlight edge information [51–53].

Medical image segmentation is usually considered a network architecture and/or data problem. One of the major barriers is the availability of large datasets with high quality segmentations. In our study, the combination of pre-training with three anatomical priors (lung, airway, vessel) on non-cancerous lungs followed by finetuning on tumour labels improved segmentation performance despite a small training dataset. It is most likely that pre-training provided additional anatomical context and created bounding regions to localise the tumour. Using anatomical priors have been previously shown to improve segmentations on heart, brain, liver, pancreas, and lymph nodes [52,54–57]. This approach has been less explored in lung tumours. Mathai et al (2025) trained two nnU-Net models, one without anatomical priors and one with 28 anatomical priors, for lung nodule segmentation [58]. Although incorporating 28 anatomical priors reduced vDSC score from 0.75 (±0.19) to 0.73 (±0.18), it managed to localise all false positives within the lung parenchyma unlike the baseline model [58].

This suggests a mechanism of constraining predictions within anatomical regions through priors [58]. However, using 28 anatomical priors may be adding too much noise. Asha et al (2024) utilised a transfer learning approach using the Segment Anything Model to leverage bounding box prompts for segmenting the LIDC-IDRI cohort (predominantly lung nodules) [59]. Although not exactly using anatomical priors, this approach achieved the best vDSC score of 0.97 compared to other DL networks [59]. It is important to note that these studies focussed primarily on lung nodules.

There remains significant interest in exploring radio-biomarkers for lung cancer as it is non-invasive and allows 3D spatial analysis of the tumour. Furthermore, compared to other-omics (e.g. genomics, transcriptomics, computational pathology) radio-biomarkers are cheaper and faster to develop for clinical use. Previous radiomic research in resectable NSCLC has primarily focused on stage 1 disease or combining stage 1-3 [25–28]. In our study, the combined clinical-radiomic model performed marginally better in predicting 2-year DFS compared to unimodal datasets but was not statistically significant. Staging remained the strongest predictor of recurrence in this cohort with a treatment interaction effect observed. Recurrence was weakly correlated with intra-tumoural ‘skewness’ and ‘cluster shade’, and peri-tumoural ‘cluster shade’. Given the weak predictive signal, these results need to be interpreted with caution. We found that most radiomic features were not robust to perturbation experiments and only 20-25% have an excellent concordance (ICC ≥0.9). Bootstrapping was wrapped around the feature selection method, mRMR-RFE-RF, to provide a realistic view of model stability and demonstrated that majority of features are not repeatable. Most radiomic studies do not integrate ICC and bootstrapping within their pipeline, which may explain why many radiomic signatures have not been reproduced. Batch effects are known to impact traditional radiomic features and this barrier will continue to exist [60,61]. Whilst older harmonisation techniques may be used, there is increasing promise in more flexible DL approaches [62].

Alternatively, developing batch-invariant radio-biomarkers is another option (e.g. shape). Traditional shape features are usually not prognostic and novel approaches need to be considered. Wu et al (2021) previously defined four prognostic radiological subtypes of NSCLC clustered based on tumour volume, regional margin variation, shape symmetry, and shape regularity [63]. Peri-tumoural vascular morphology is another promising but under investigated radio-biomarker [64]. The emerging role of anti-angiogenic drugs in both AGA and non-AGA advanced NSCLCs highlight the importance of this mechanism. The use of deep features has also been increasing. For example, deep features derived from 3D CNN in stage 1-3 NSCLCs was correlated with genetic markers of cellular proliferation [65]. Radio-biomarkers, however, cannot fully encapsulate the complex tumour microenvironment compared to histopathology and genomics. In the era of AI, integration of multi-modal datasets will be the future. However, the role of radio-biomarkers remains to be defined. It is likely that traditional radiomics is limited by the voxel resolution of clinical-grade CTs. The use of high-resolution imaging, such as micro-CT, has yet to be explored. Although micro-CT is currently limited to human ex-vivo scanning, it may have a role in post-operative risk-stratification in early-stage NSCLCs.

Our study has several limitations. First, although our sample size compared favourably with the literature for this type of lung cancer cohort, it remains small and potentially underpowered. Second, we excluded death from the primary endpoint. Due to high missingness in cause of death we could not accurately identify cancer-specific mortality. By excluding these patients, this reduced our sample size. Third, batch effects could have impacted radiomic stability. Although ComBat harmonisation is available, it requires the assumption that batches have similar biological variance and that differences are driven by technical factors [66]. In this cohort, due to the range of tumour shapes and invasiveness, ComBat harmonisation could mask biological signals. To adjust for this, we explicitly evaluated radiomic repeatability through these variations using perturbations and bootstrapping. Finally, there was significant treatment heterogeneity with regards to whether a patient received SACT and the type of SACT given. Although SACT was not significantly associated with recurrence in our study, several phase 3 randomised control trials have proven its benefit. These differences could represent the underpowering of our study. However, a stage-dependent effect was recognised during interaction analysis.

## 5.0 Conclusion

In summary, we developed a semi-automated pipeline incorporating deep learning auto-segmentation to explore robust radiomic biomarkers in resected stage 2A-3B NSCLC. Our findings show that auto-segmentation can be effectively integrated within radio-biomarker discovery with an acceptable error margin. Although it is possible to identify robust radiomic biomarkers using this approach, the overall predictive signal remains weak and the majority of radiomic features lack reproducibility. The development of batch-invariant radio-biomarkers is vital for the consideration of future clinical adoption within oncology. Integrating auto-segmentation pipelines during biomarker developments will make these approaches clinically scalable.

## Data Availability

Due to the potentially identifiable nature of the raw data regarding our patients, this information will remain confidential and not shared. The TCIA datasets and nnU-Net are open-source, and can be found on the following websites: https://www.cancerimagingarchive.net/ & https://github.com/MIC-DKFZ/nnUNet.

