## Supplemental Figures for "A Deep Learning Lung Cancer Segmentation Pipeline to Facilitate CT-based Radiomics"

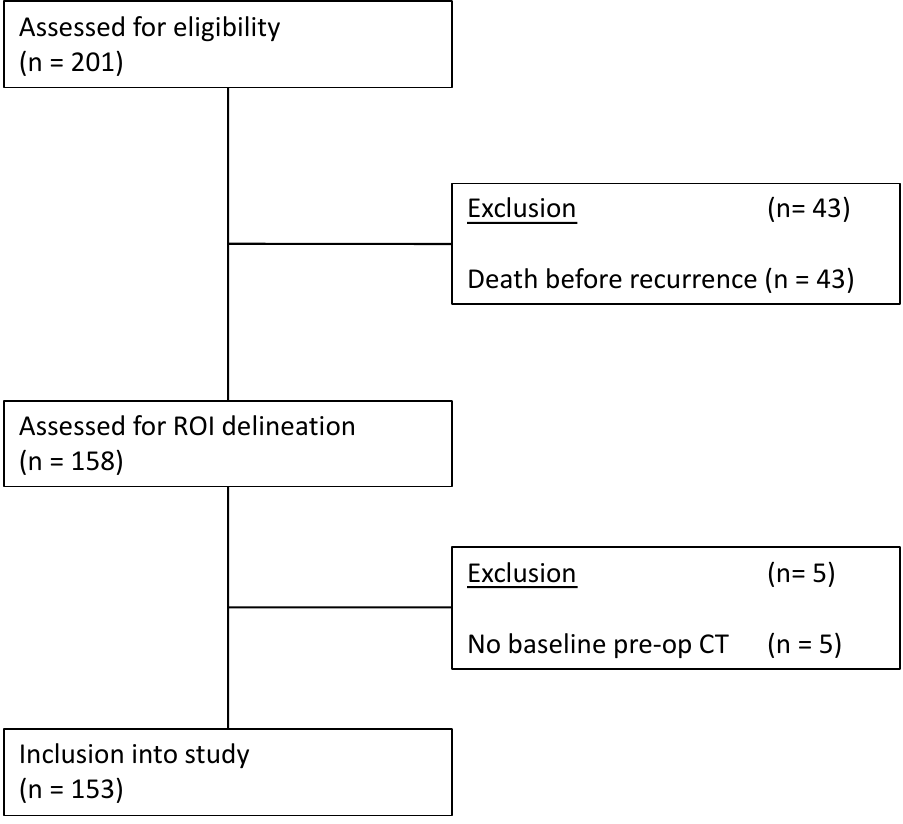

**Supplemental Fig. 1:** CONSORT statement of patient selection. ROI, region of interest.

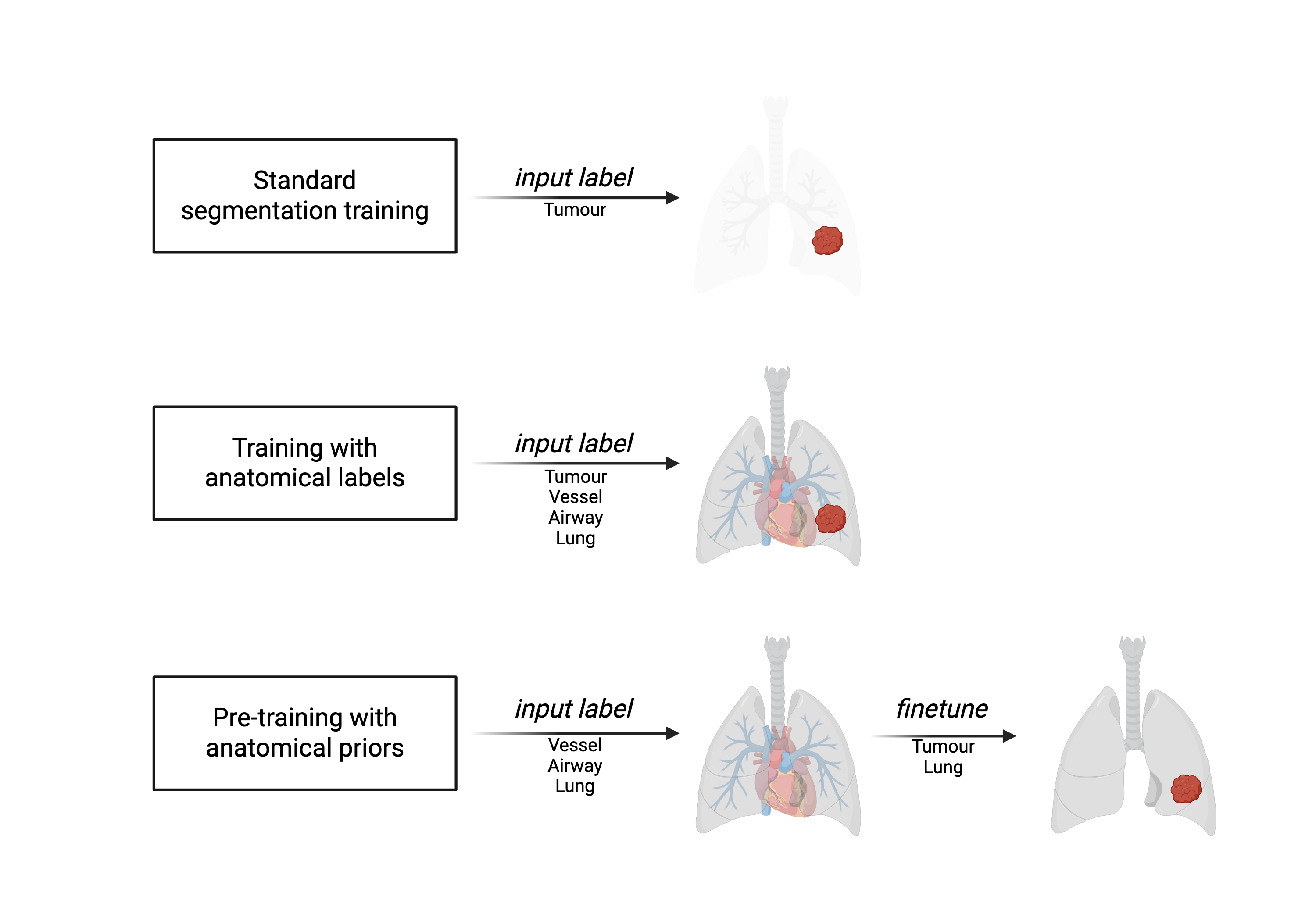

**Supplemental Fig. 2:** Different approached to nnU-Net training. Standard segmentation training involves inputting tumour-label alone. Training with anatomical labels involves inputting lung, airway, and vessel labels and training along with tumour-labels. Hierarchy of training is as ordered lung (4), airway (3), vessel (2), and tumour (1). Pre-training involves initially training the nnU-Net with non-cancer containing lungs with anatomical labels: lung (3), airway (2), vessel (1). After training, the weights are re-used and finetuned on cancer-containing cases with lung (2) and tumour (1) labels. Figure created with BioRender.

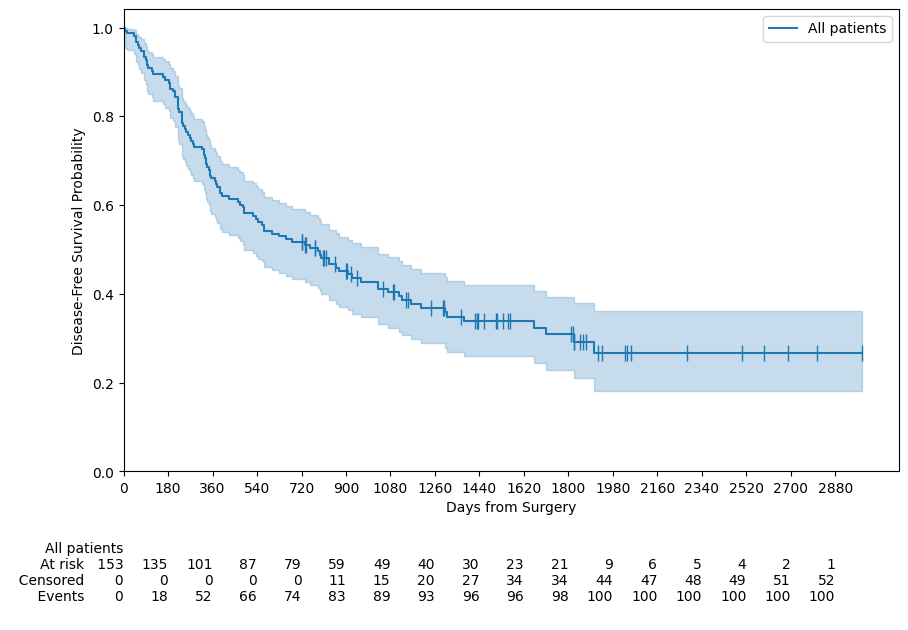

**Supplemental Figure 3:** Kaplan-Meier survival curve on disease-free survival. Vertical lines denote censored events. Blue shaded region represents 95%CI.

| **Case 1:** excellent segmentation  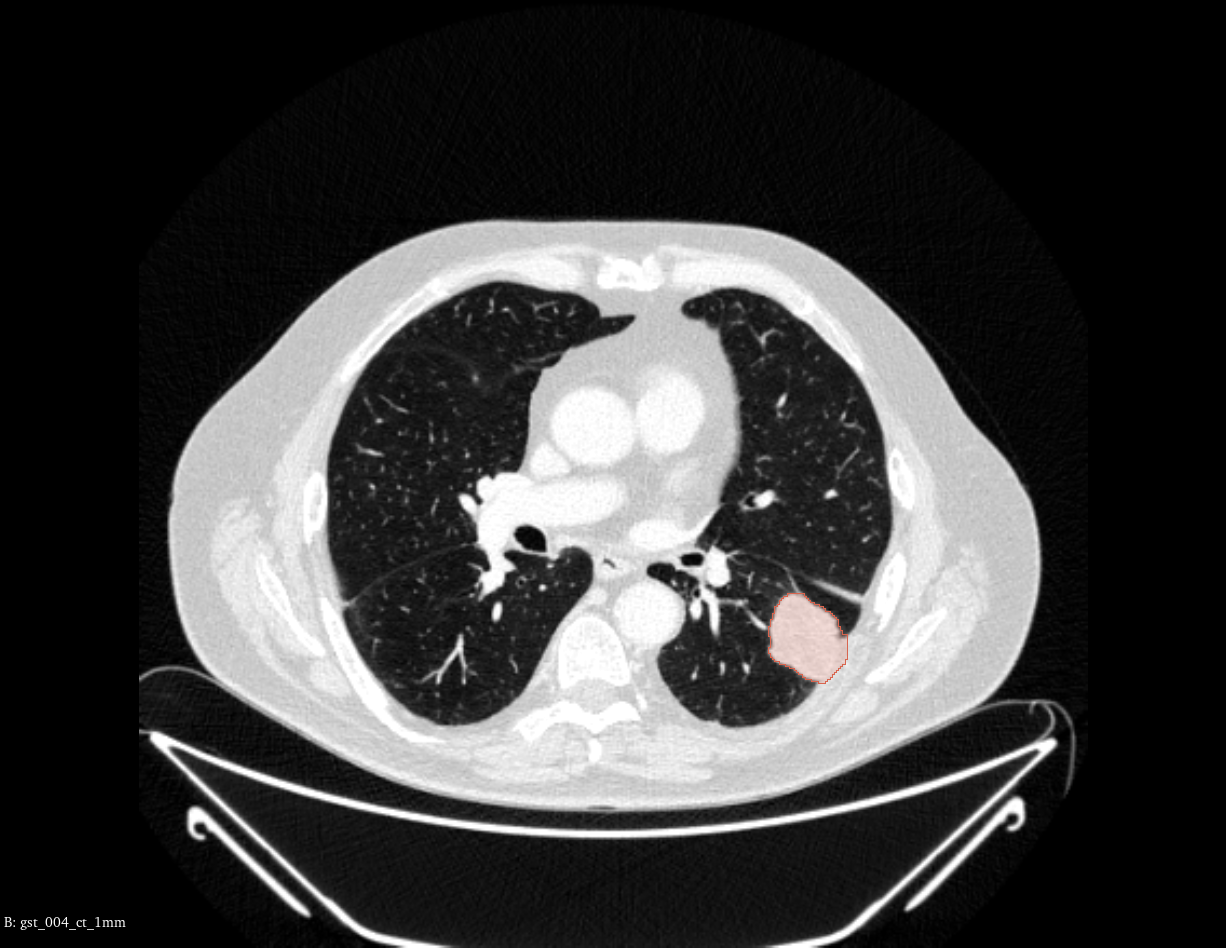 | **Case 2:** excellent segmentation  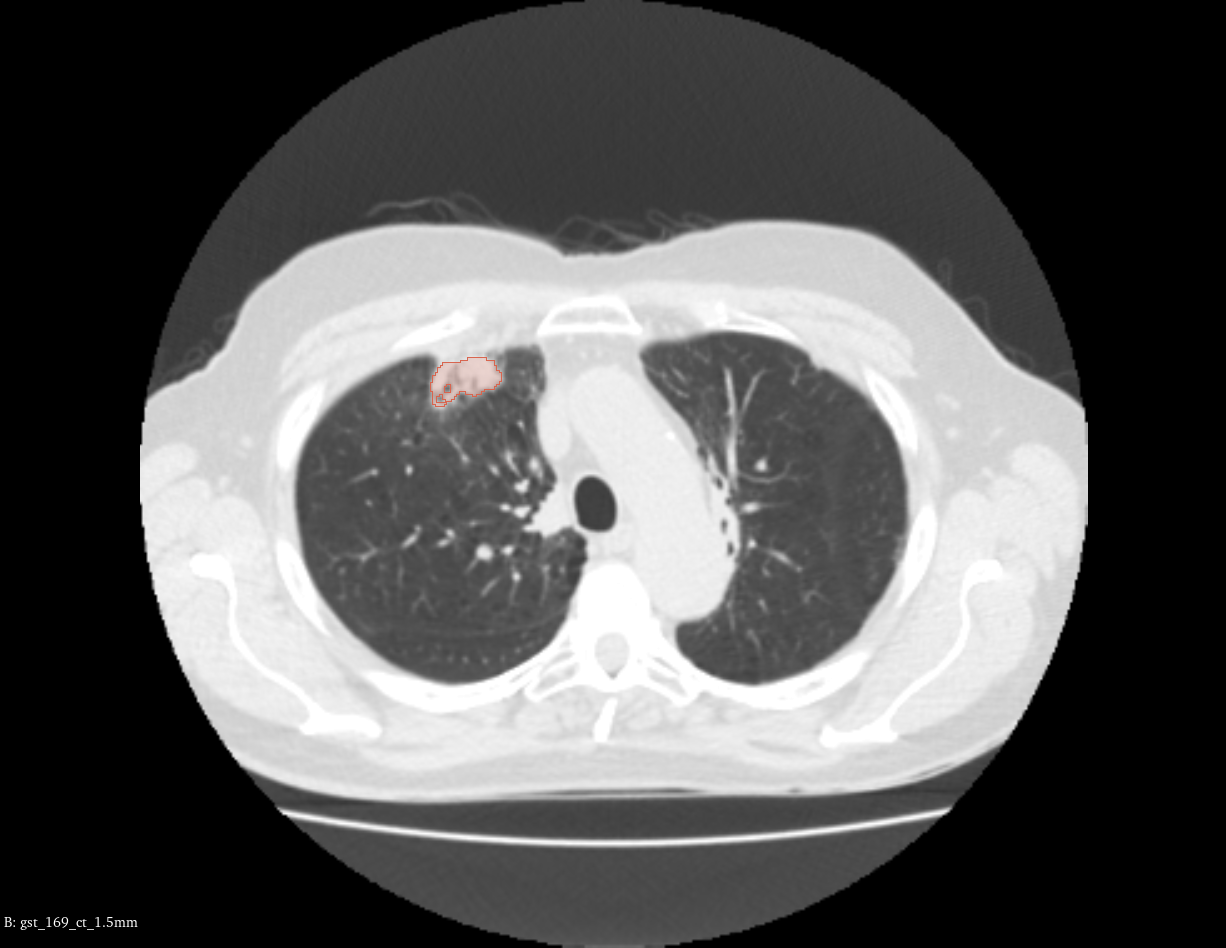 | **Case 3:** excellent segmentation  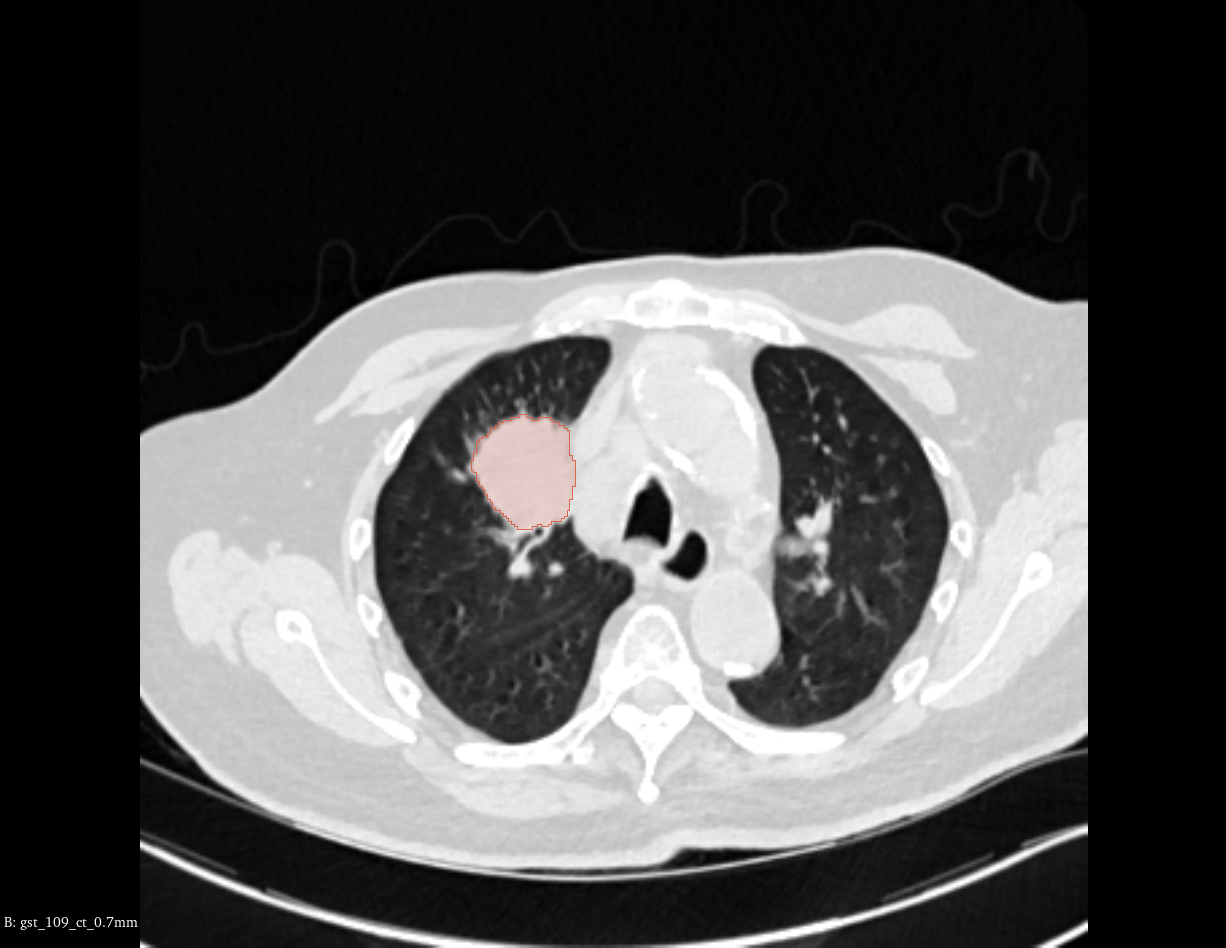 |
| --- | --- | --- |
| **Case 4:** minor corrections  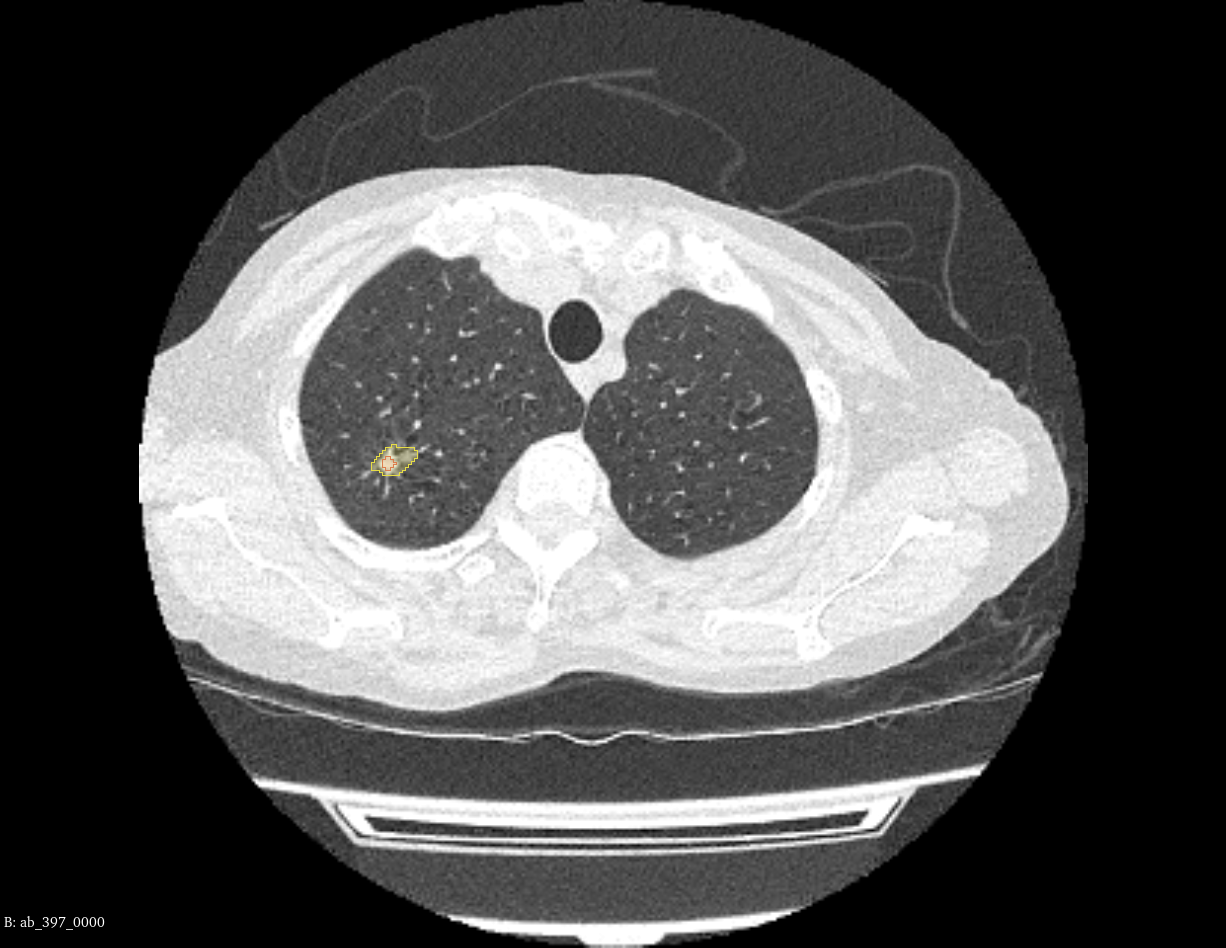 | **Case 5:** minor corrections  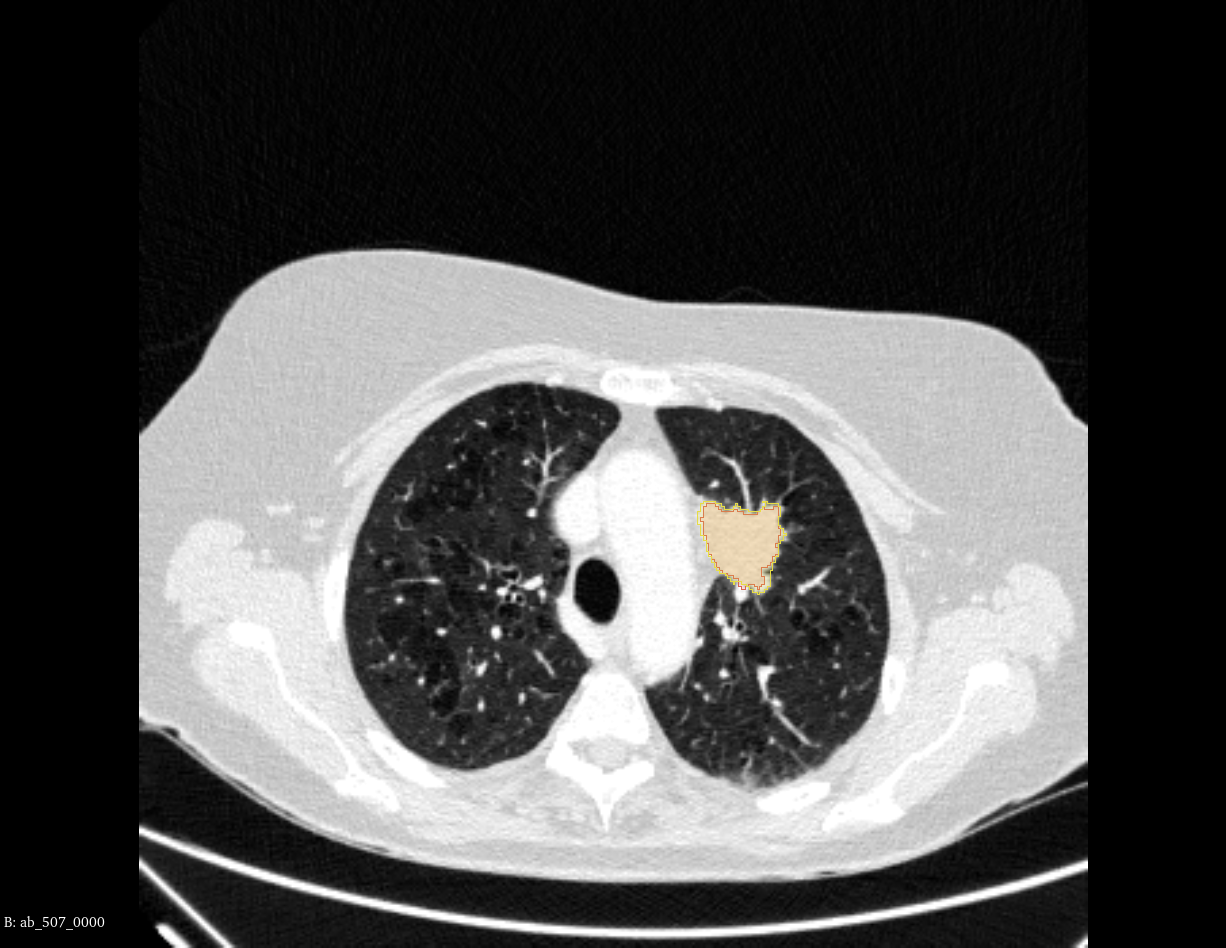 | **Case 6:** minor corrections  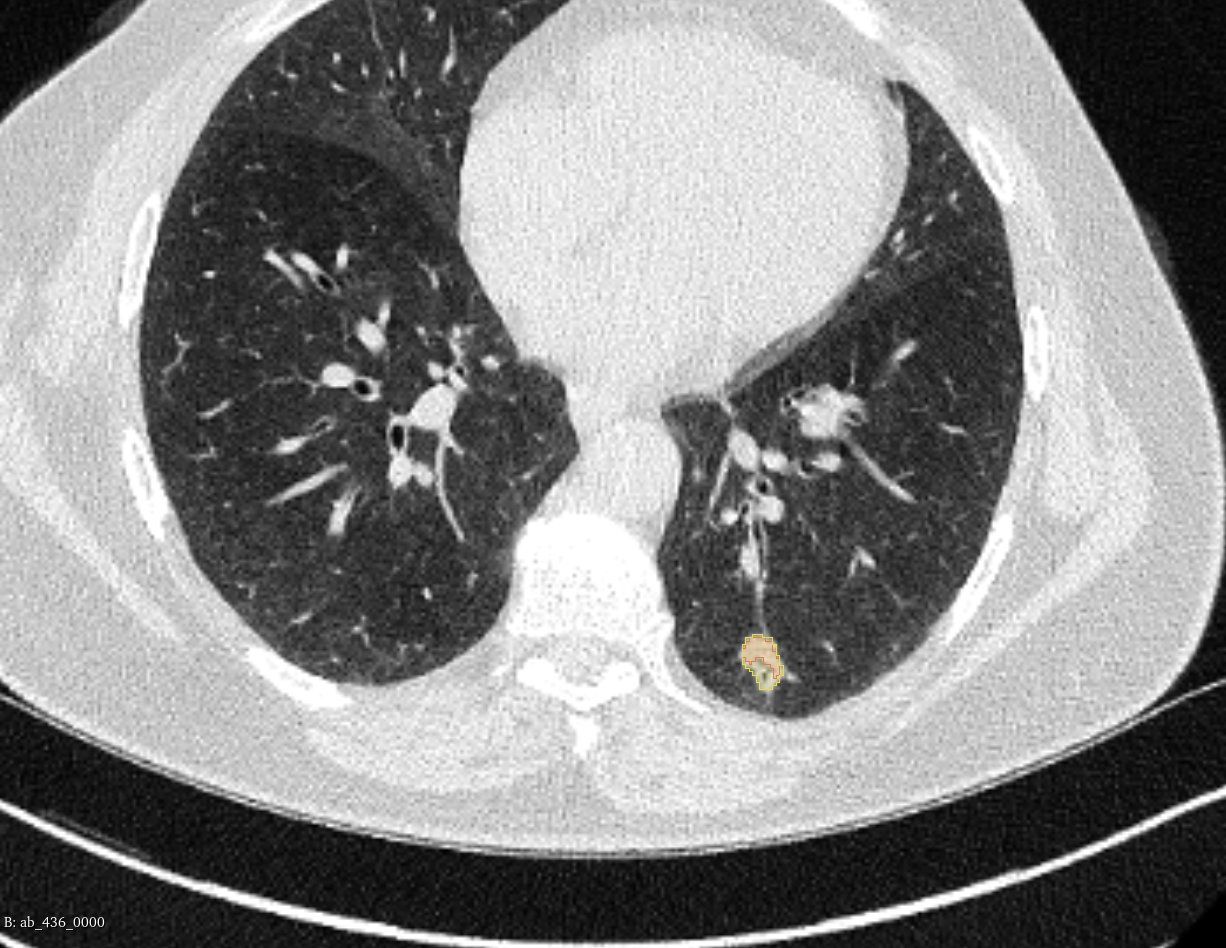 |
| **Case 7:** major corrections  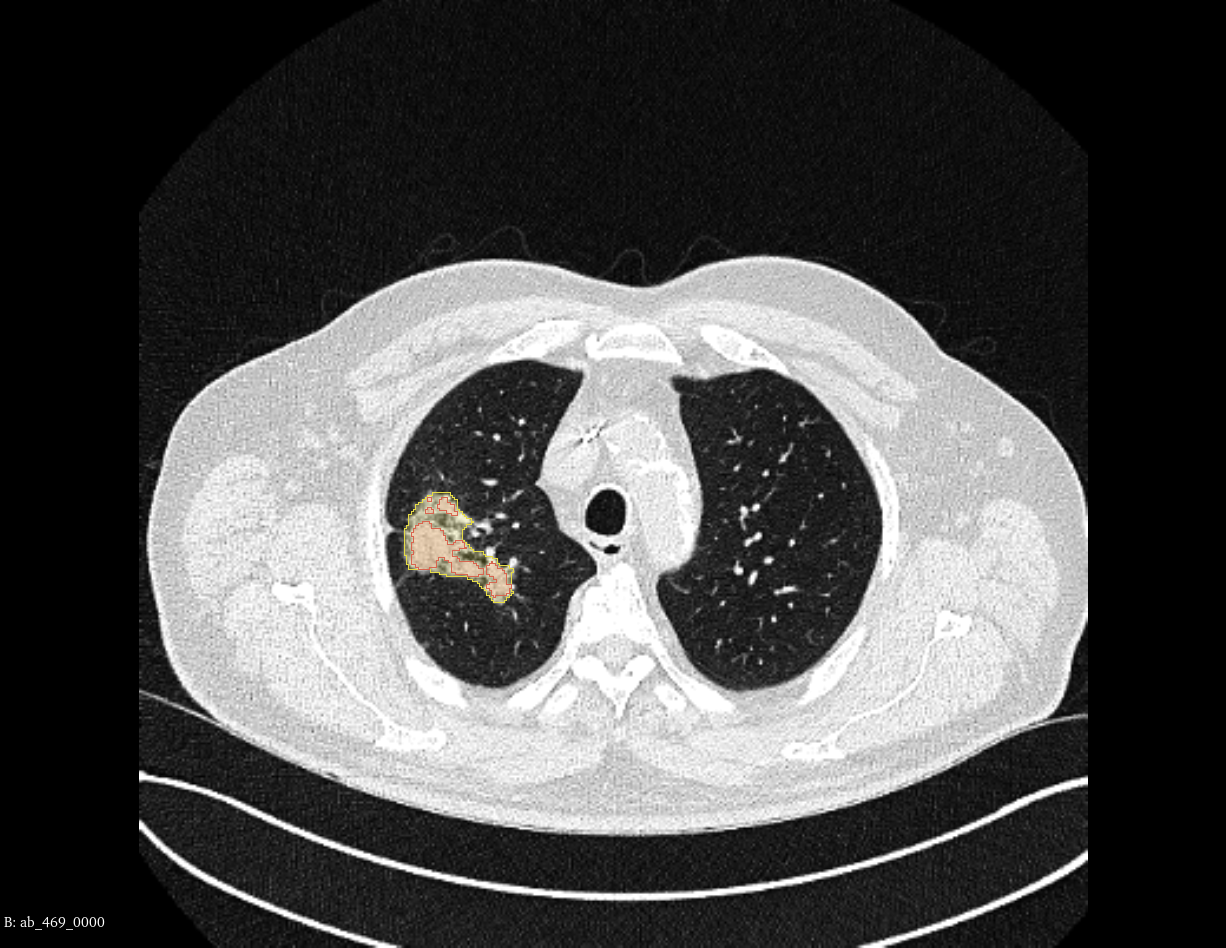 | **Case 8:** major corrections  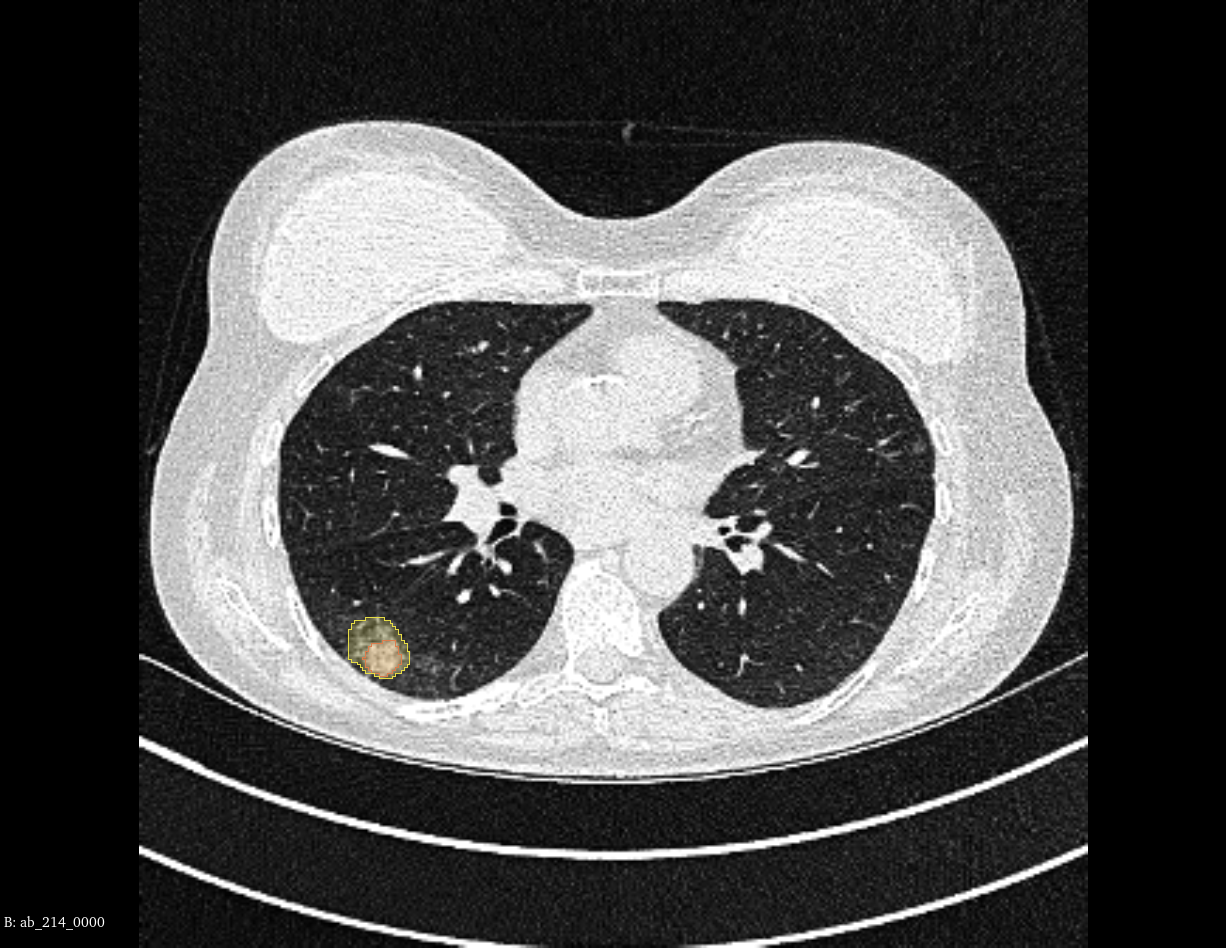 | **Case 9:** major corrections  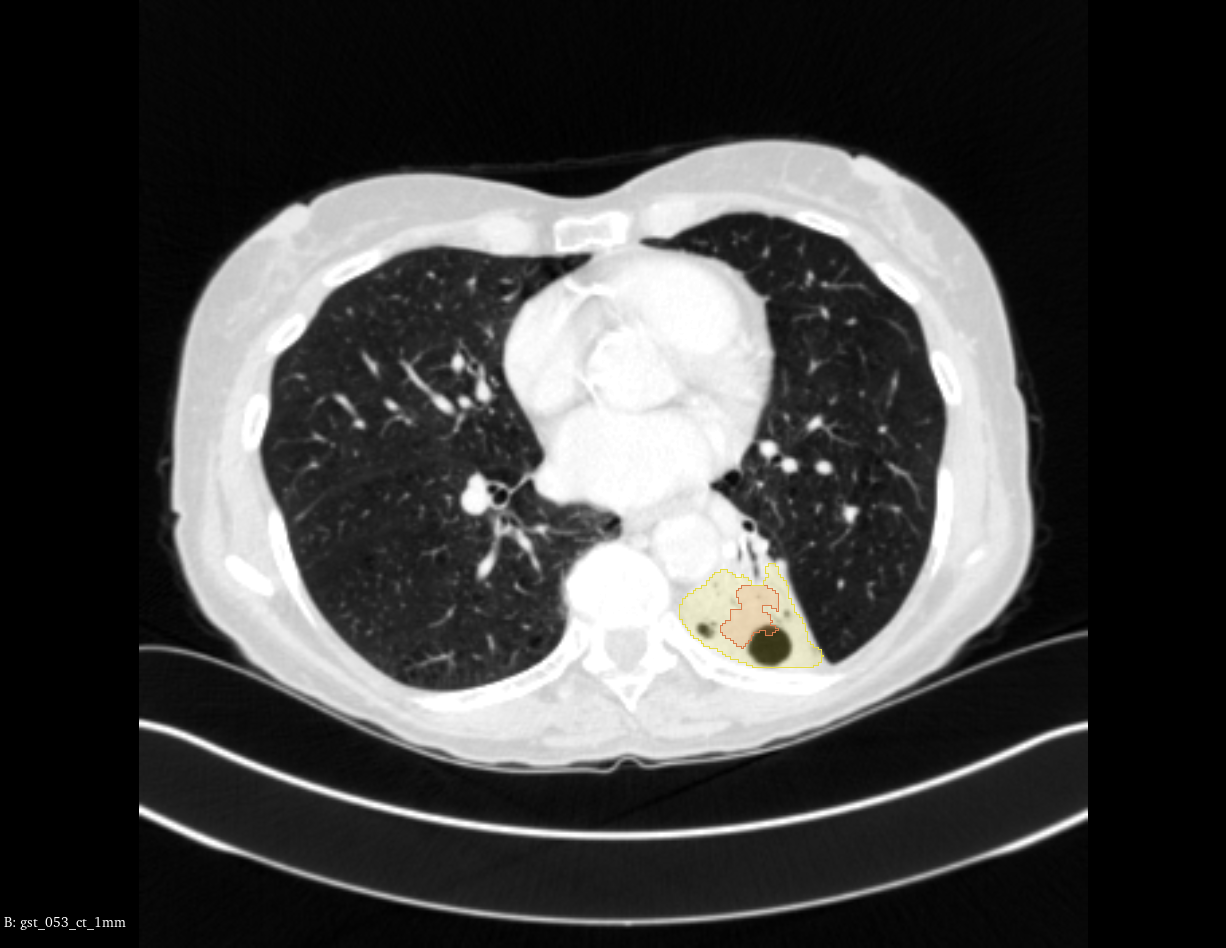 |
| **Case 10:** missed segmentation  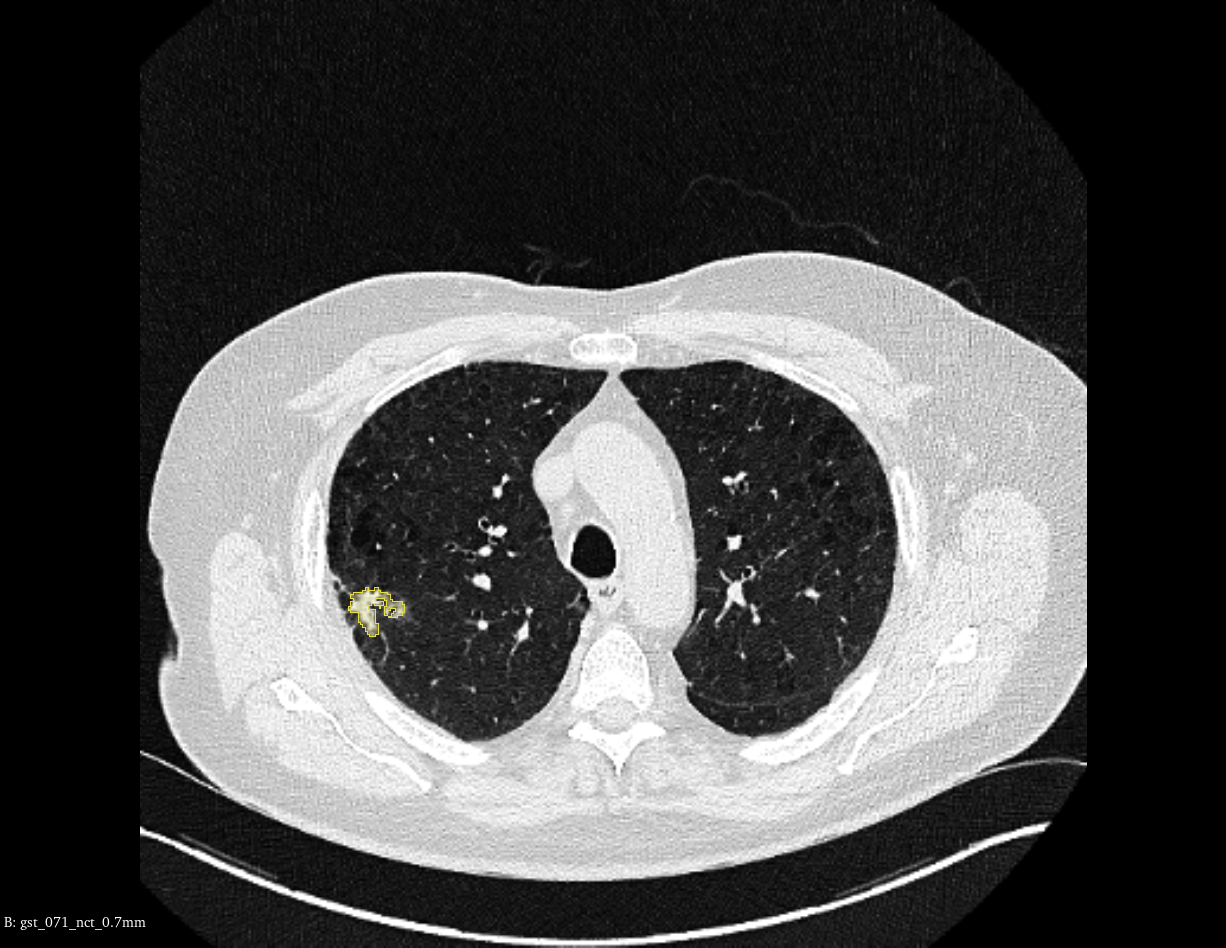 | **Case 11:** missed segmentation  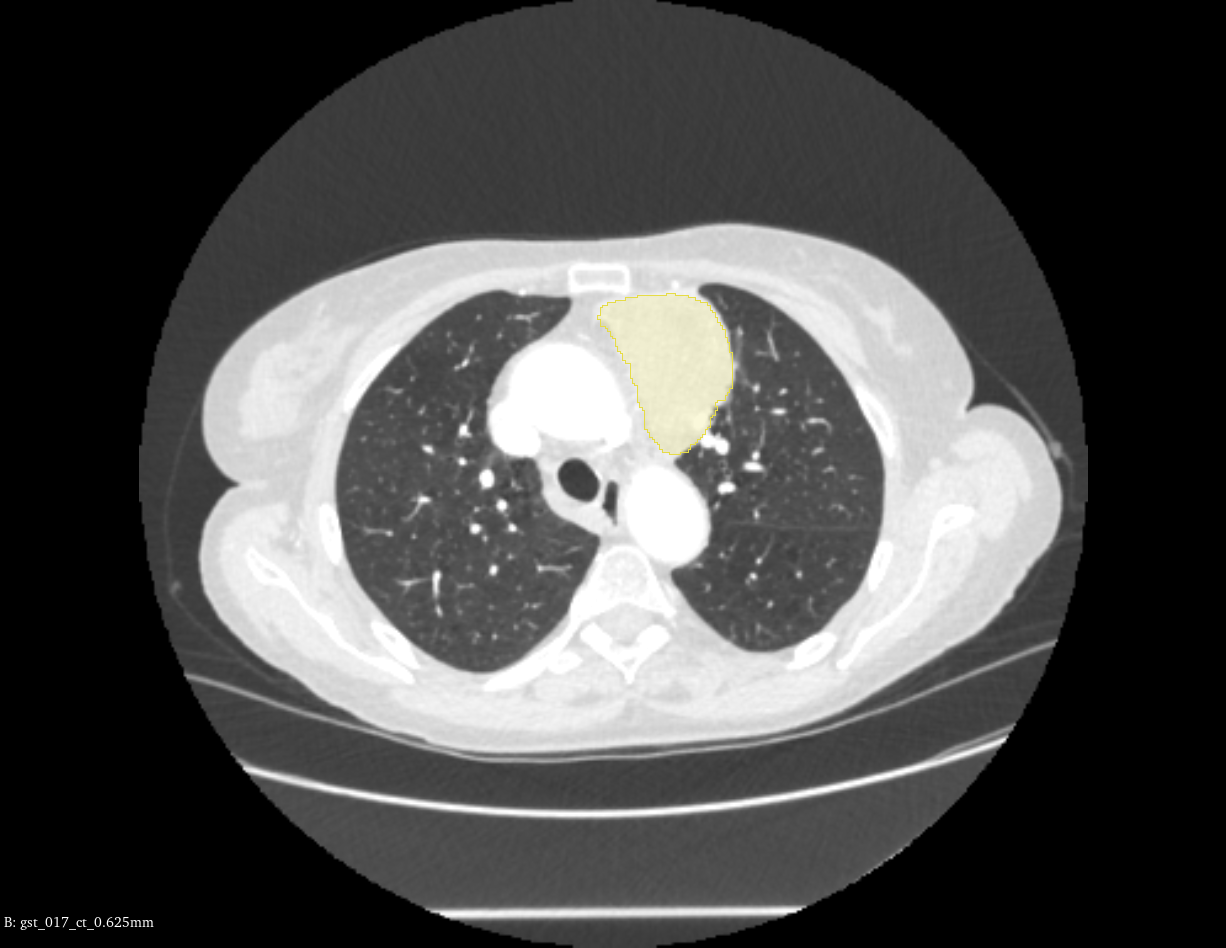 | **Case 12:** missed segmentation  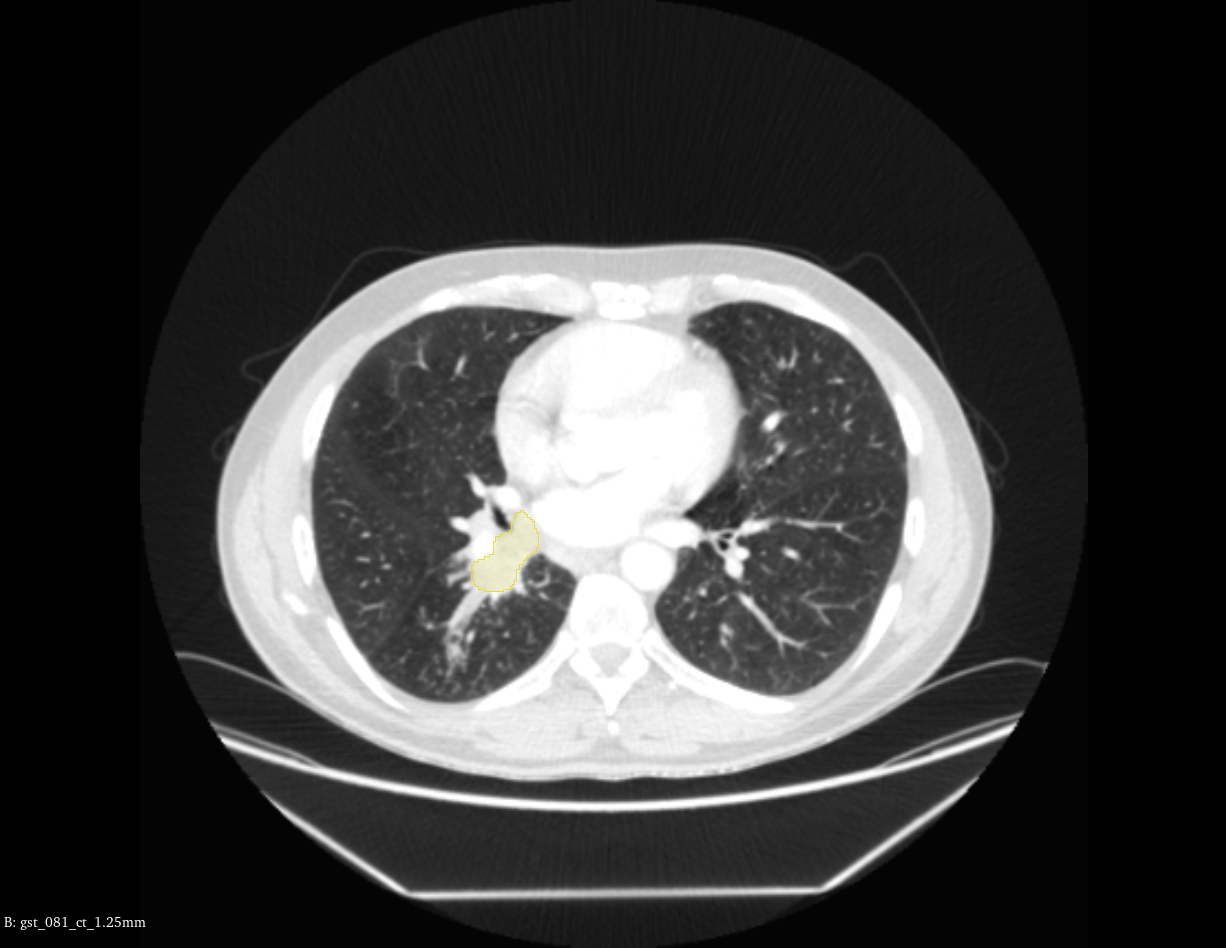 |

**Supplemental Figure 4:** Visual examples of cases requiring no amendments (case 1-3), minor changes (case 4-6), major changes (7-9), and missed segmentations (case 10-12) from our pretrained auto-segmentation model. Red segmentation = nnU-Net auto-segmentation. Yellow segmentation = ground truth segmentation.

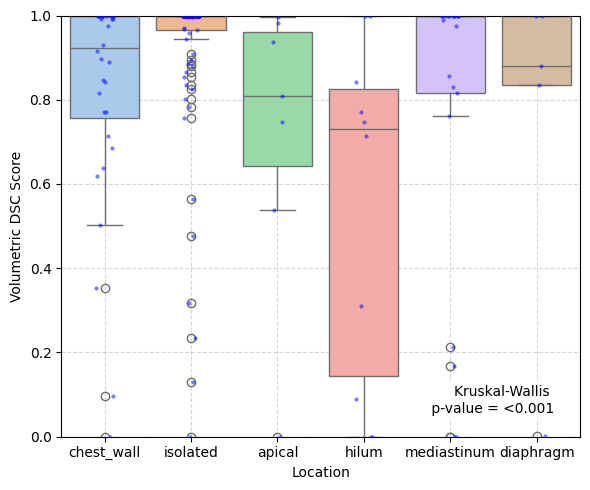

**Supplemental Figure 5A:** Comparison of volumetric DSC scores with tumour location. Isolated tumours refer to tumours located within the lung parenchyma that is not in contact with other structures.

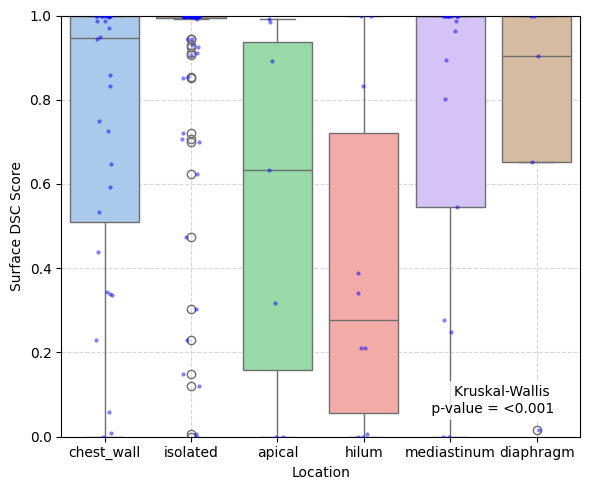

**Supplemental Figure 5B:** Comparison of surface DSC scores with tumour location. Isolated tumours refer to tumours located within the lung parenchyma that is not in contact with other structures.

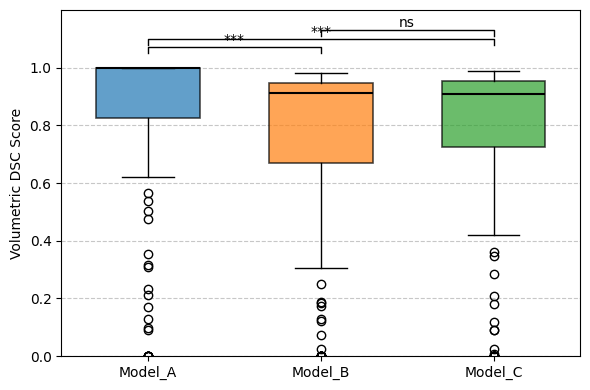

**Supplemental Figure 6A:** Box-whiskers plot comparing volumetric DSC scores across different models. ***, *p*-value <0.001. ns, not significant (*p*-value > 0.05).

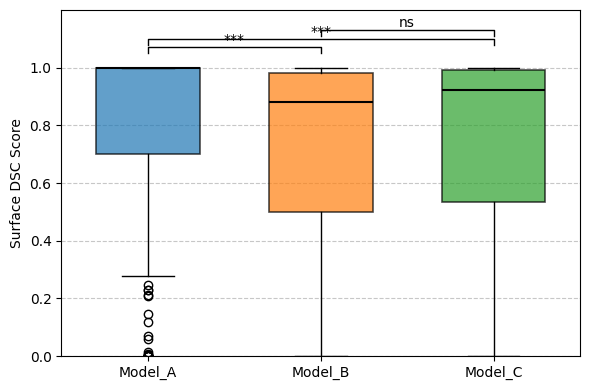

**Supplemental Figure 6B:** Box-whiskers plot comparing surface DSC scores across different models. ***, *p*-value <0.001. ns, not significant (*p*-value > 0.05).

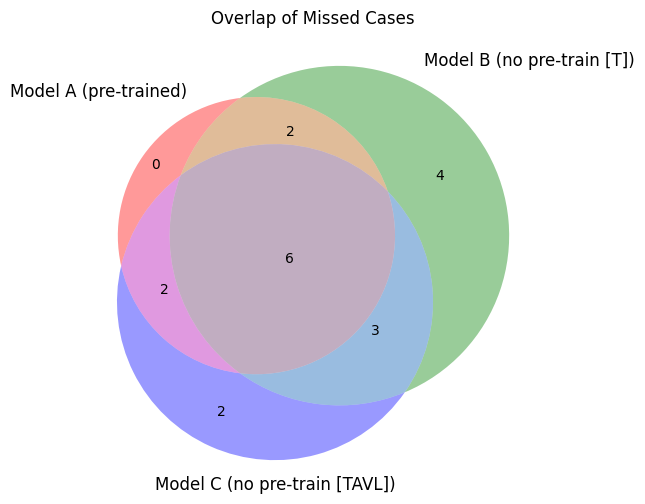

**Supplemental Figure 7A:** Venn diagram showing overlap of missed auto-segmentations across different models.

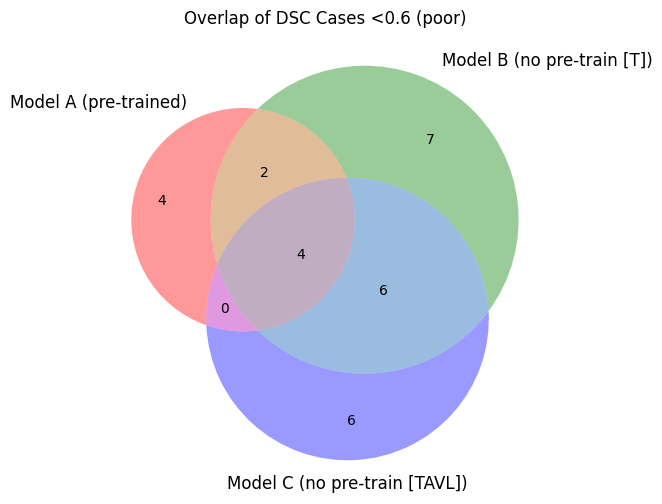

**Supplemental Figure 7B:** Venn diagram showing overlap of poor auto-segmentations (DSC <0.6; requiring major changes) across different models.

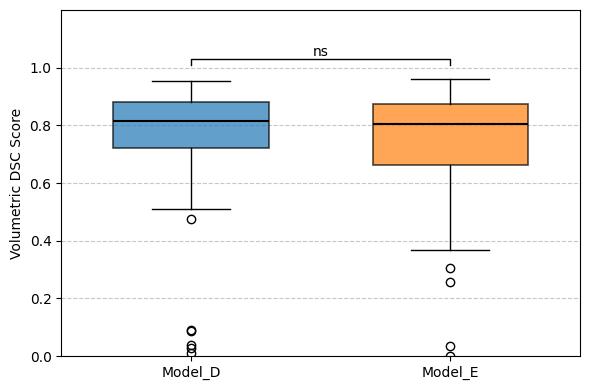

**Supplemental Figure 8A:** Box-whiskers plot comparing volumetric DSC scores across different models on reverse validation. ns, not significant (*p*-value > 0.05).

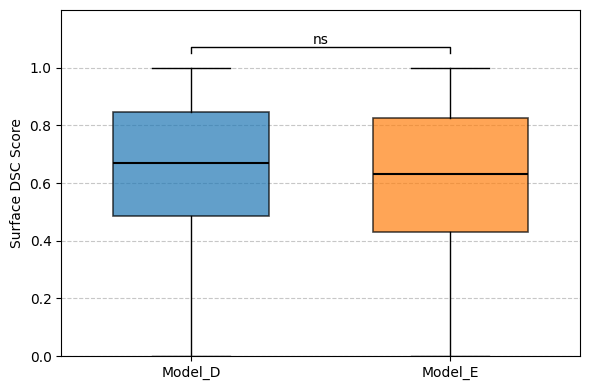

**Supplemental Figure 8B:** Box-whiskers plot comparing surface DSC scores across different models on reverse validation. ns, not significant (*p*-value > 0.05).

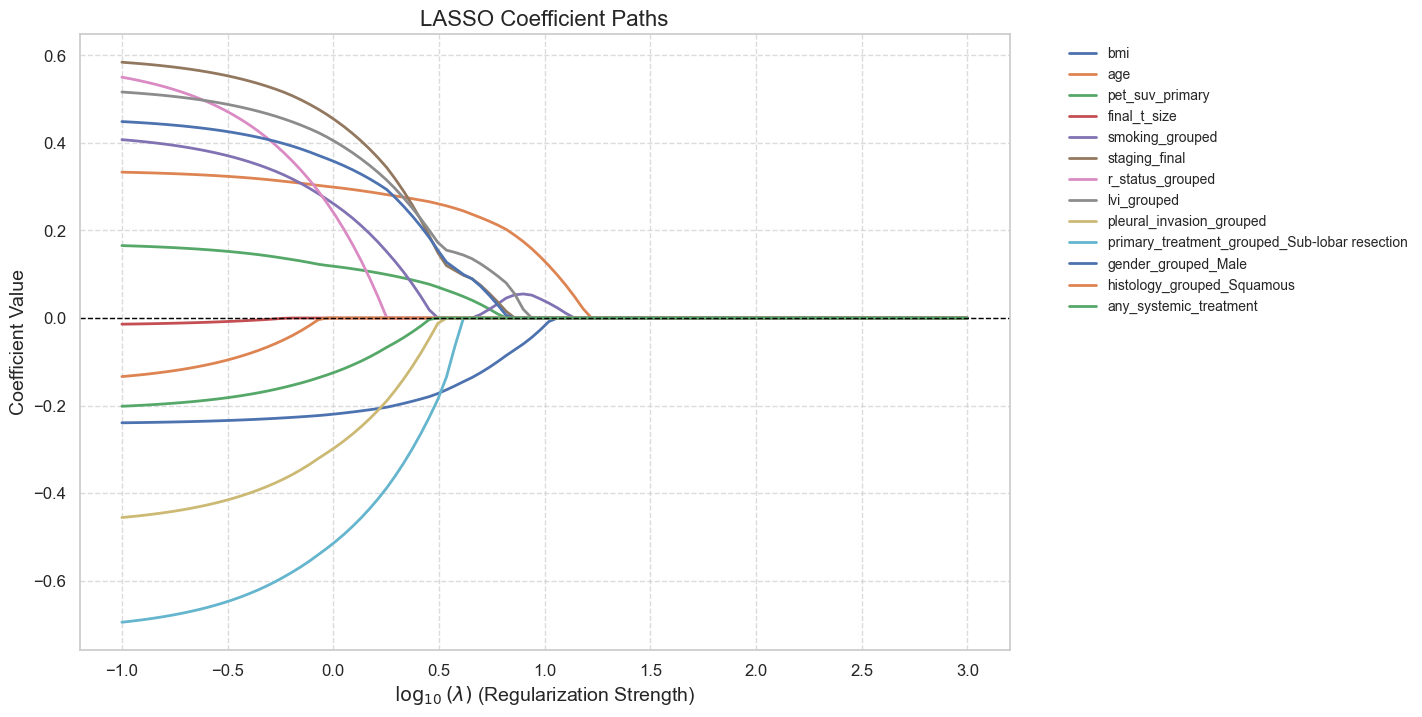

**Supplemental Figure 9:** LASSO coefficient path plot for clinical feature selection.

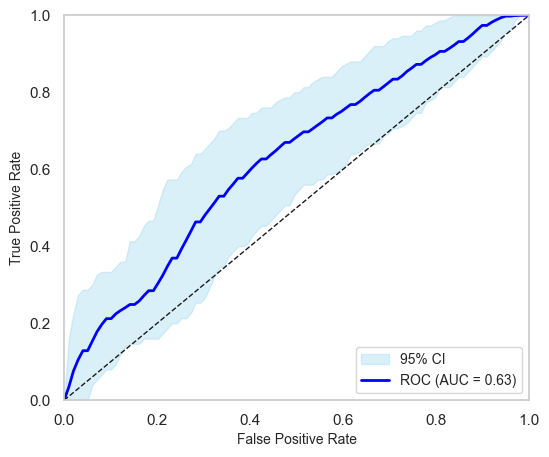

**Supplemental Figure 10A:** ROC-AUC curve of classic approach in clinical feature selection.

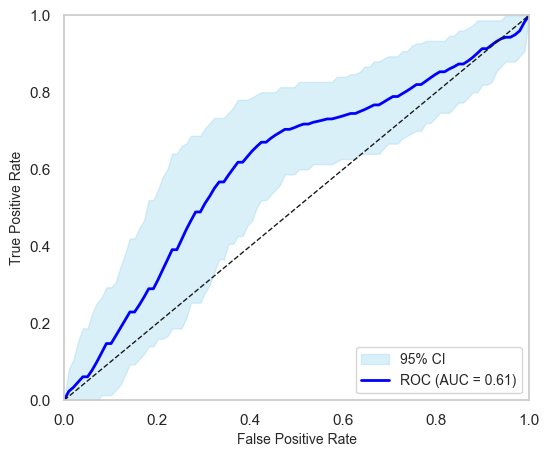

**Supplemental Figure 10B:** ROC-AUC curve of LASSO-selected clinical features.

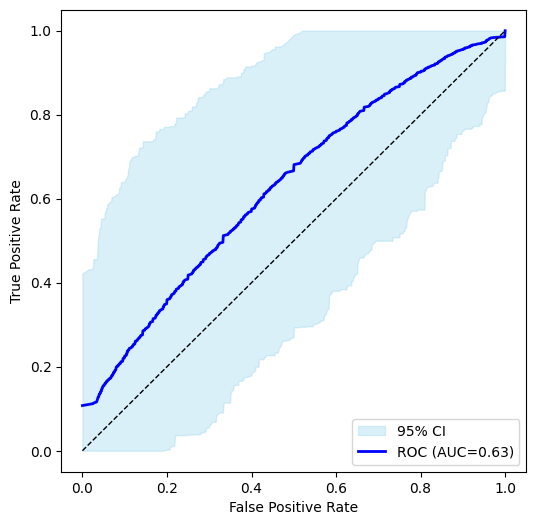

**Supplemental Figure 11A:** ROC-AUC curve of intra-tumoural-only radiomics model.

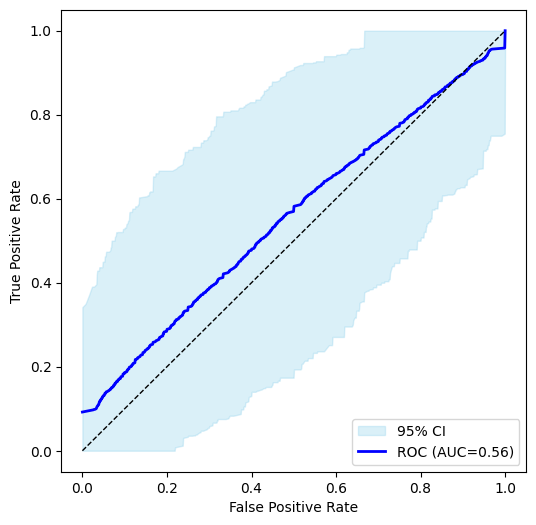

**Supplemental Figure 11B:** ROC-AUC curve of peri-tumoural-only radiomics model.

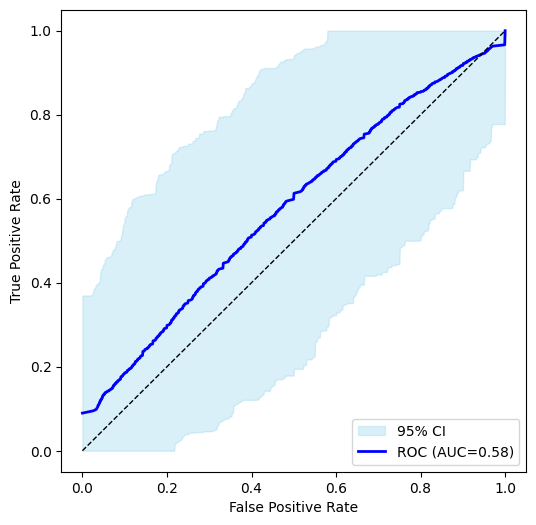

**Supplemental Figure 11C:** ROC-AUC curve of whole lung-only radiomics model.

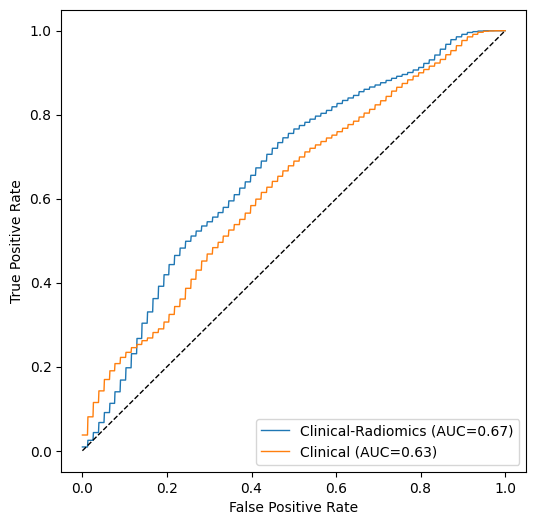

**Supplemental Figure 12:** ROC-AUC curve of clinical-radiomic versus clinical-alone model.

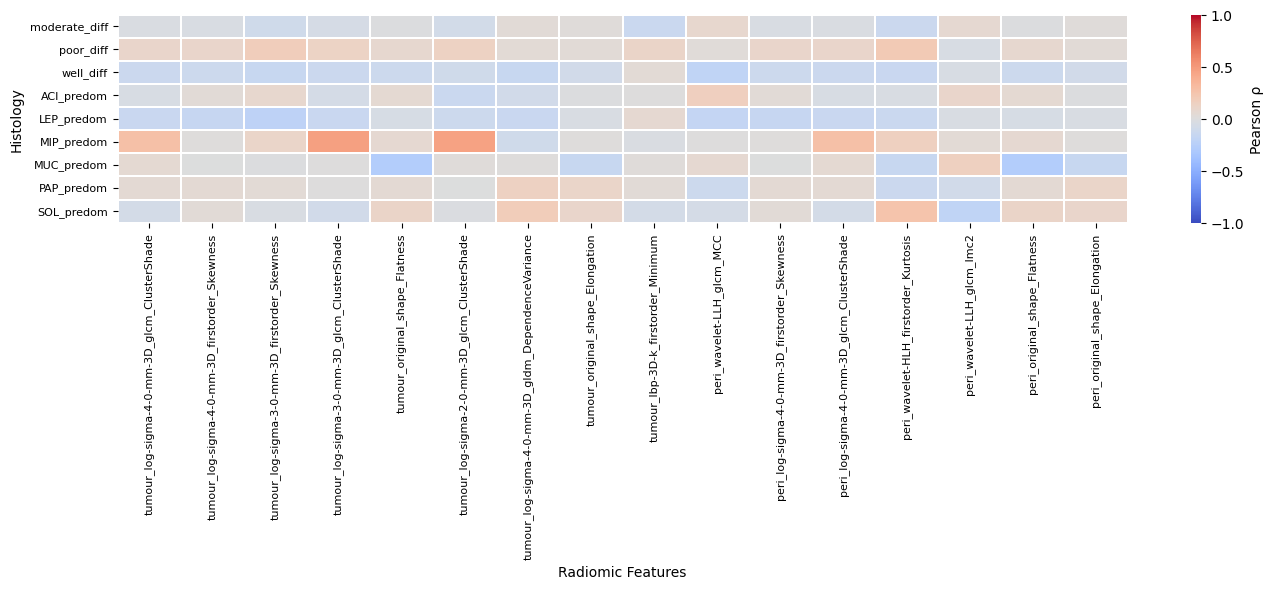

**Supplemental Figure 13:** Pearson correlation matrix of histological grading (*n*=75/153) and predominant growth pattern (*n*=71/153) with radiomic features. Prefix of radiomic features indicate which region-of-interest it was derived from: tumour_ = intra-tumoural; peri_ = peri-tumoural; lung_ = whole lung.
