## Supplemental Tables for "A Deep Learning Lung Cancer Segmentation Pipeline to Facilitate CT-based Radiomics"

| Manufacturer | Protocol | Convolution kernel | Slice thickness (mm) | Pixel size (mm) | Tube energy (kVp) | Tube current  (mA) | Collimation single width (mm) |
| --- | --- | --- | --- | --- | --- | --- | --- |
| Philips:  23.5%  GE:  25.5%  SIEMENS:  29.4%  Canon:  0.6%  Toshiba:  20.9 | Chest with contrast:  47.7%  Chest without contrast:  28.8%  High-resolution CT Chest:  2.6%  Chest-Abdomen-Pelvis with contrast:  20.9% | Smooth:  42.5%  Sharp:  57.5% | Mean:  1.2  Median:  1.0  Range:  0.5-5.0 | Mean:  0.76  Median:  0.75  Range:  0.46-0.98 | Mean:  113  Median:  120  Range:  90-120 | Mean:  202  Median:  173  Range:  30-649 | Mean:  0.60  Median:  0.60  Range:  0.50-1.25 |
| Missing:  0% | Missing:  0% | Missing:  0% | Missing:  0% | Missing:  0% | Missing:  0% | Missing: 10.5% | Missing:  38.6% |

**Supplemental Table 1:** CT imaging and scanner metadata.

|  | Age (years) | Sex | No lung abnormality | COPD (GOLD stage) | ILD |
| --- | --- | --- | --- | --- | --- |
| Summary | Median 66 (range 58-74) | Male: 65.2% | 21.7% | GOLD 2: 34.8% GOLD 3: 17.4%  GOLD 4: 17.4% | 21.7% |

**Supplemental Table 2**: Summary of pre-training characteristics from the SUMMIT dataset (n=23). SUMMIT is a UK-based national lung cancer screening trial using low-dose CT. COPD, chronic obstructive pulmonary disease. GOLD, Global Initiative for COPD. ILD, interstitial lung disease.

|  | Age (years) | Sex | Stage | Histology |
| --- | --- | --- | --- | --- |
| RIDER LUNG CT (*n*=8) [46]* | Mean 62.1 | Male: 50% | Unknown – mixture of primary and metastatic lesions | Unknown (NSCLC) |
| NSCLC-Radiomics-Interobserver1 (*n*=8) [47] | Mean 60.5 | Male: 50% | Stage 1: 25%  Stage 2: 12.5%  Stage 3: 62.5% | LUAD: 50%  LUSC: 12.5%  NOS: 25%  NEC: 12.5% |
| NSCLC-Radiomics (*n*=10) [48] | Mean 69.9 | Male: 50% | Stage 1: 30% Stage 2: 20%  Stage 3: 50% | LUAD: 30%  LUSC: 30%  NOS: 20%  NEC: 20% |
| NSCLC-Radiogenomics (*n*=12) [49] | Mean 68.2 | Male: 75% | Stage 1: 25%  Stage 2: 41.7%  Stage 3: 25%  Stage 4: 8.3% | LUAD: 58.3%  LUSC: 33.3%  NOS: 8.3% |
| LUAD-CT-SURVIVAL (*n*=12) [50]* | Mean 67 | Male: 47.5% | Stage 1: 35%  Stage 2: 25%  Stage 3: 32.5%  Stage 4: 7.5% | LUAD: 100% |

**Supplemental Table 3:** Summary of training characteristics from the five TCIA dataset (n=50). *Patient level data was unavailable and therefore total cohort baseline characteristics are presented. LUAD, lung adenocarcinoma. LUSC, lung squamous cell carcinoma. NOS, not otherwise specified. NEC, neuroendocrine carcinoma. NSCLC, non-small cell lung cancer. TCIA, The Cancer Imaging Archive.

|  | Description |
| --- | --- |
| Missed cases  (*n*=10/153) | - flat opacity with irregular margins, abuts chest wall - irregular opacity, abuts mediastinum, presence of prominent apical cap - large mass invading mediastinum and chest wall - irregular subpleural consolidation with background interstitial lung disease and structural distortion of background lung - irregular central opacity with bronchial obstruction - irregular nodule - solid cavitating lesion, abuts chest wall - irregular intraluminal tumour - irregular nodule adjacent to interlobar fissure, abuts pleura - nodule abuts pulmonary vein |
| Cases requiring major changes  (*n*=10/153) | - semi-solid nodule with cystic changes and surrounding ground-glass opacities (suspicious for multifocal pre-invasive disease) - multiple pulmonary nodules - irregular apical mass - predominantly solid mass with air bronchogram within it, surrounding ground-glass opacities - multiple irregular pulmonary nodules - irregular mass with air bronchogram within it, multiple satellite nodules - large cavitating mass with heterogenous contrast enhancement - prominent soft tissue density surrounding hilum - multiple ground glass opacities with semi-solid lesion - large consolidation with central necrosis and associated volume loss of surrounding lung |

**Supplemental Table 4:** Description of missed auto-segmentation cases and cases requiring major change from auto-segmentation model.

|  | TCIA  n=50 (%) | GCC  n=153 (%) |
| --- | --- | --- |
| Location |  |  |
| Isolated | 28 (56) | 78 (50.98) |
| Chest wall | 12 (24) | 32 (20.92) |
| Mediastinum | 4 (8) | 21 (13.73) |
| Hilum | 4 (8) | 10 (6.54) |
| Apical | 2 (4) | 7 (4.58) |
| Diaphragm | 0 | 5 (3.27) |
| Tumour type |  |  |
| Solid | 47 (94) | 126 (82.35) |
| Semi-solid | 3 (6) | 27 (17.65) |
| Volume (mm^3^) |  |  |
| Mean (±std) | 31802.20 (± 52349.72) | 21003.13 (±37647.64) |
| Median (range) | 9229 (215-271414) | 10727 (118-34658) |
| Diameter (mm) |  |  |
| Mean (±std) | 49.64 (±25.25) | 67.55 (±59.89) |
| Median (range) | 45.53 (10.25-121.70) | 49.35 (8.77-326.83) |
| Sphericity (0-1) |  |  |
| Mean (±std) | 0.64 (±0.11) | 0.69 (±0.12) |
| Median (range) | 0.64 (0.29-0.83) | 0.72 (0.39-0.86) |

**Supplemental Table 5:** Comparison of TCIA (training) and GCC (external validation) datasets regarding radiological phenotypes. Isolated tumour location refers to tumour not adhering to any major boundary structures or hilum. The remaining locations refer to tumour adherence to that location (e.g. chest wall refers to tumour adherence to the chest wall, indicating no visible lung parenchyma on CT between the two edges). Diameter measured by maximum 3D diameter. Sphericity as extracted from PyRadiomics, the closer the value to 1 the more spherical the shape. Brackets denoting percentages refers to within column percentages. GCC, Guy’s Cancer Cohort. Std, standard deviation. TCIA, The Cancer Imaging Archive.

|  | Volumetric DSC scores | | | Surface DSC scores | | |
| --- | --- | --- | --- | --- | --- | --- |
|  | Spearman *ρ* | U-statistic | *p*-value | Spearman *ρ* | U-statistic | *p*-value |
| Volume | -0.21 | – | 0.009 | -0.30 | – | <0.001 |
| Diameter | -0.28 | – | <0.001 | -0.33 | – | <0.001 |
| Sphericity | 0.60 | – | <0.001 | 0.63 | – | <0.001 |
| S:V ratio | -0.03 | – | 0.691 | 0.04 | – | 0.592 |
| Contrast | – | 3936 | <0.001 | – | 3347 | <0.001 |
| Kernel | – | 2253 | <0.001 | – | 3926 | <0.001 |
| Pixel size | <0.01 | – | 0.994 | -0.02 | – | 0.780 |
| Slice thickness | -0.04 | – | 0.662 | 0.03 | – | 0.726 |

**Supplemental Table 6:** Univariate analysis of relationship between shape and batch factors with DSC scores. Spearman rank correlation was used on continuous variables, whilst Mann-Whitney U test was used on categorical variables. The direction of difference in the Mann-Whitney U test is: presence of IV contrast > no contrast; smooth kernel > sharp kernel.

|  | Model_D | Model_E | Model_D vs E | |
| --- | --- | --- | --- | --- |
|  | Score (±std) | Score (±std) | z-score | *p*-value |
| vDSC | 0.73 (±0.25) | 0.72 (±0.23) | 1.04 | 0.304 |
| sDSC | 0.64 (±0.26) | 0.62 (±0.26) | 0.92 | 0.363 |
| IoU | 0.62 (±0.24) | 0.60 (±0.24) | 0.94 | 0.349 |
| Sensitivity | 0.77 (±0.25) | 0.77 (±0.24) | 0.19 | 0.851 |
| Specificity | 1.00 (±<0.01) | 1.00 (±<0.01) | -0.13 | 0.890 |
| Precision | 0.79 (±0.24) | 0.74 (±0.26) | 0.41 | 0.688 |
| Accuracy | 1.00 (±<0.01) | 1.00 (±<0.01) | -0.19 | 0.848 |

**Supplemental Table 7:** Wilcoxon-rank pairwise comparison of auto-segmentation metrics across models during reverse validation. Model_D: pre-trained using anatomical priors (lung, airway, vessel) on cases without lung tumours, then finetuned on cases with lung cancers. Model_E: no pre-training, trained on tumour labels as well as anatomical labels (lung, airway, vessel). DSC, Dice-Sørensen coefficient. IoU, intersection over union. sDSC, surface DSC. std, standard deviation. vDSC, volumetric DSC.

|  | Univariate Analysis | | Multivariate Analysis | |
| --- | --- | --- | --- | --- |
|  | OR (95%CI) | *p*-value | OR (95%CI) | *p*-value |
| Age (years) | 1.28 (0.92–1.77) | 0.14 | – | – |
| Sex (male vs female) | 2.16 (1.13–4.14) | 0.02 | 2.05 (1.02–4.11) | 0.04 |
| Smoking history (current vs ex vs never) | 1.24 (0.72–2.13) | 0.44 | – | – |
| BMI | 0.83 (0.60–1.14) | 0.25 | – | – |
| Histology (squamous vs non-squamous) | 0.98 (0.47–2.06) | 0.96 | – | – |
| Stage (3 vs 2) | 2.09 (1.09–4.00) | 0.03 | 2.25 (1.10–4.61) | 0.03 |
| Primary tumour size (mm) | 1.20 (0.87–1.66) | 0.26 | – | – |
| PET-SUV max of primary tumour | 1.22 (0.88–1.69) | 0.23 | 1.22 (0.86–1.71) | 0.27 |
| EGFR/KRAS status  WT vs EGFR mutated  KRAS mutated vs EGFR mutated  Unknown vs EGFR mutated | 1.65 (0.86–3.14)  0.98 (0.46–2.08)  0.53 (0.25–1.16) | 0.13  0.95  0.11 | –  –  – | –  –  – |
| Primary surgery (sub-lobar vs lobectomy) | 0.39 (0.13–1.18) | 0.09 | 0.43 (0.13–1.39) | 0.16 |
| Adjuvant or Neoadjuvant treatment (yes or no) | 0.98 (0.52–1.86) | 0.95 | 0.79 (0.39–1.60) | 0.52 |
| Pleural invasion (PL1-2 vs PL0) | 0.82 (0.43–1.56) | 0.54 | 0.67 (0.33–1.35) | 0.26 |
| LVI (yes vs no) | 1.94 (1.01–3.72) | 0.05 | – | – |
| R status (R1-2 vs R0) | 2.85 (0.85–9.51) | 0.09 | 2.07 (0.58–7.45) | 0.26 |

**Supplementary Table 8:** Univariate and multivariate analysis on original candidate clinical features. Multivariate analysis results only displayed for final selected clinical features.

|  | AUC | Accuracy | F1 score | Sensitivity | Specificity | Brier score |
| --- | --- | --- | --- | --- | --- | --- |
| Classic selection | 0.63 (0.54-0.71) | 0.59 (0.51-0.66) | 0.54 (0.44-0.63) | 0.49 (0.37-0.60) | 0.68 (0.58-0.78) | 0.24 (0.22-0.27) |
| LASSO selection | 0.61 (0.52–0.70) | 0.63 (0.56–0.71) | 0.63 (0.55-0.70) | 0.64 (0.53-0.75) | 0.63 (0.53-0.74) | 0.25 (0.22-0.28) |

**Supplemental Table 9:** Multivariate logistic regression model performance of clinical features selected from the classic approach (staging, R status, sex, PET-SUV-max, reception of SACT, pleural invasion, type of primary surgical treatment) versus LASSO feature reduction (staging, sex, primary tumour diameter, T stage, PET-SUV-max, age, BMI). Although primary tumour diameter and T stage were selected by LASSO, T2-T4 can be staged independently due to other factors such as presence of pleural invasion, local invasion, and satellite nodules. Brackets denote 95%CI.

| Region-of-interest | Feature | Filter |
| --- | --- | --- |
| Intra-tumoural | shape_Flatness  shape_Elongation  firstorder_Skewness  firstorder_Minimum  gldm_DependenceVariance  glcm_ClusterShade | original  original  log_sigma_3-0-mm-3D / 4-0-mm  lbp-3D-k  wavelet-LHH  log-sigma-2-0-mm-3D / 3-0-mm / 4-0-mm |
| Peri-tumoural | shape_Flatness  shape_Elongation  firstorder_Skewness  firstorder_Kurtosis  glcm_MCC  glcm_ClusterShade  glcm_Imc2 | original  original  log-sigma-4-0-mm-3D  wavelet-HLH  wavelet-LLH  log-sigma-4-0-mm-3D  wavelet-LLH |
| Whole lung | firstorder_Mean  glcm_MCC | wavelet-HHL  wavelet-HHL |

**Supplemental Table 10:** Most common radiomic features selected across ≥50% bootstraps.
